# High prevalence of female genital schistosomiasis and under-detection by urine microscopy among women of reproductive age in Kilifi County, Kenya

**DOI:** 10.64898/2026.04.01.26349935

**Authors:** Hellen Wambui Kariuki, Sophia Mogere Nyasore, Felistas Wayua Muthini, Peter Waweru Mwangi, Patrick Makazi, Marianne Wanjiru Mureithi, Wallace Bulimo, Eric Wanjala, Frank Onyambu, Lyle Mckinnon, Kariuki Njaanake

## Abstract

**Background:** Female genital schistosomiasis (FGS) is a neglected gynaecological manifestation of *Schistosoma haematobium* (*S. haematobium*) infection, resulting from the deposition of parasite eggs in the female genital tract. Although urogenital schistosomiasis is highly prevalent in parts of coastal Kenya, including Kilifi County, the burden of FGS among women of reproductive age remains poorly characterised. Routine diagnosis of *S. haematobium* infection relies largely on urine microscopy, which may underestimate genital involvement. This study aimed to assess the prevalence, diagnostic concordance, and risk factors for FGS among women of reproductive age in Kilifi County, Kenya.

**Methodology:** In this cross-sectional study, 320 randomly selected women aged 15–50 years were recruited from rural Kilifi County; 261 provided complete data for analysis. A structured questionnaire was administered to collect sociodemographic and behavioural information. Urinary schistosomiasis was assessed using triplicate urine microscopy over three consecutive days, and FGS was evaluated using real-time polymerase chain reaction (PCR) targeting the *S. haematobium Dra1* gene sequence on self-collected high vaginal swabs.

**Results:** Overall, the prevalence of PCR-confirmed FGS was 36.0% (94/261), while urinary egg excretion was detected in 13.0% (34/261) of participants. Concordance between urine microscopy and genital PCR was 70.9%. Notably, 72% of women with PCR-confirmed FGS had no detectable parasite eggs in their urine. In bivariate analyses, factors such as urinary infection severity, water contact behaviours, haematuria, dysuria, age group, place of residence, and prior history of schistosomiasis were found to be associated with female genital schistosomiasis (FGS). However, in the multivariable logistic regression, only sub-location and urinary infection severity remained independently associated with the infection. Additionally, PCR cycle threshold (Ct) values showed a non-linear relationship with mean urinary egg counts, indicating that the detection of genital parasite DNA does not directly correspond to the urinary egg burden.

**Conclusion:** FGS prevalence among women in Kilifi County was substantially higher than indicated by urine microscopy alone. The majority of women with genital schistosomiasis did not exhibit detectable urinary egg excretion, highlighting the limitations of routine parasitological screening for identifying genital disease. These findings underscore the need to incorporate genital sampling and molecular diagnostics into schistosomiasis control strategies targeting women of reproductive age in endemic settings.

**Author’s summary:** Female genital schistosomiasis (FGS) is a neglected gynaecological manifestation of *Schistosoma haematobium* infection, occurring when parasite eggs lodge in the female genital tract. Although schistosomiasis control programmes typically rely on urine microscopy to detect infection, this approach may miss genital disease. FGS has been linked to chronic inflammation and adverse reproductive health outcomes; yet its burden among adult women in endemic areas remains poorly documented.

In this study, we investigated FGS among women of reproductive age in Kilifi County, coastal Kenya, using molecular testing of self-collected genital swabs. We found that more than one-third of women had evidence of FGS. Notably, nearly three-quarters of women with genital infection had no detectable parasite eggs in their urine. This finding indicates that routine urine-based screening alone may substantially underestimate the true burden of genital schistosomiasis in endemic communities.

Our results highlight a hidden burden of FGS among women of reproductive age and underscore the need to strengthen diagnostic approaches within schistosomiasis control programmes. Integrating genital sampling, improving surveillance, and reinforcing water, sanitation, and hygiene (WASH) interventions, alongside mass drug administration, could enhance early detection and reduce ongoing transmission, ultimately improving women’s reproductive health in endemic settings.

## INTRODUCTION

Female genital schistosomiasis (FGS) is a neglected gynaecological disease caused by the deposition of *Schistosoma haematobium* (*S. haematobium*) in the female reproductive tract (1). Approximately 30-56 million women and girls, mainly in sub-Saharan Africa, are affected by FGS (2). In Kenya, *S. haematobium* is mainly endemic to the coastal region (3–5). A hospital-based study in Kwale County reported a 23% prevalence of *S. haematobium* infection, with genital pathology observed in 46.7% of the infected women (6). FGS is characterised by genital lesions, unusual vaginal discharge, pelvic pain, vaginal bleeding, dysmenorrhoea, dyspareunia and granulomatous inflammation (7–13). If left untreated, it can lead to adverse reproductive sequelae including Human Immunodeficiency Virus/Sexually Transmitted Infections (HIV/STIs) acquisition, spontaneous abortion, ectopic pregnancy, subfertility and cervical cancer (14–17). Women with FGS commonly experience stigma, shame, psychological distress and intimate partner violence due to the symptoms and complications it manifests (17,18).

*Bulinus* species (spp.) snails are the intermediate hosts of the parasite, which produce and release cercariae into freshwater sources (19). The cercariae infect humans, the definitive host, through skin penetration upon water contact (1,20). The adult male and female worms pair up and migrate to the pelvic venous plexus, where the female lays eggs (8,21). Approximately half of the eggs manage to penetrate into the lumen of the urinary tract and are voided in urine, while the rest are trapped in various tissues in the pelvic region, invoking inflammatory responses characterised by peri-oval granulomas (22,23). In women, the ecto-endocervical junction is the most common site of egg deposition, followed by the vagina, ovaries, fallopian tubes, vulva and uterus (24–31).

In most endemic regions, urine microscopy is mainly used to diagnose *S. haematobium* infection. However, FGS can occur independent of *S. haematobium* egg excretion in urine, which can lead to misdiagnosis and under-treatment when urine microscopy is employed (32,33). Therefore, FGS is typically diagnosed by colposcopy or Polymerase Chain Reaction (PCR) (2,9,34). PCR detection of *S. haematobium* deoxyribonucleic acid (DNA), specifically targeting the *Dra1* gene, is highly sensitive and specific (35). It also enables the diagnosis of all women, including pregnant and virgin women, for whom colposcopy may be challenging.

Currently, there is no data on the prevalence of FGS in Kilifi County despite it being a major *S. haematobium* transmission zone (36). Additionally, the under-detection rate of FGS by conventional microscopy in this area is unknown. Thus, this study aimed to determine the prevalence of FGS using molecular diagnostics and to identify associated risk factors among women of reproductive age in Kilifi County.

## METHODOLOGY

### Study Design, Setting, and Ethical Considerations

This study represents the second phase of a cross-sectional investigation into the prevalence of *S. haematobium* infection and Female Genital Schistosomiasis (FGS) among women of reproductive age in rural Kilifi County, Kenya. The first phase of the study evaluated urogenital schistosomiasis (UGS) through urine microscopy within the same population from which participants for the current FGS analysis were selected. Details of the Phase I study have been documented elsewhere. The current phase focused on detecting FGS using molecular diagnostics and assessing the associated risk factors. The research was conducted in the Rabai and Magarini sub-counties, which have previously been identified as endemic for *S. haematobium*.

### Phase I: Community-based recruitment and urinary screening

In Phase I, 336 freely consenting women aged 15–50 years, long-term residents of the study area, were recruited through community-based household sampling for urinary schistosomiasis screening. Villages and sub-locations were selected from known transmission settings. In Rabai sub-county, recruitment was conducted in Ruruma Ward (Jimba and Mleji sub-locations). In Magarini sub-county, recruitment was undertaken in Magarini Ward (Pumwani and Marikebuni sub-locations), Garashi Ward (Bore, Singwanya, Masindeni, and Mikuyuni sub-locations), and Sabaki Ward (Sabaki sub-location along the Athi–Galana–Sabaki River) (**Figure 1**). Participants were selected using proportional allocation across villages based on the number of women of reproductive age. Eligible households were visited systematically using a predetermined sampling interval. Only one eligible woman per household was enrolled, and where more than one eligible woman was present, simple random (lottery) selection was applied to minimise intra-household clustering. The planned minimum sample size was approximately 150 women per sub-county (total ≈300), calculated using single-proportion estimation with conservative prevalence assumptions, adjusted for clustering and anticipated non-response. Recruitment yielded 336 participants.

**Figure 1.**
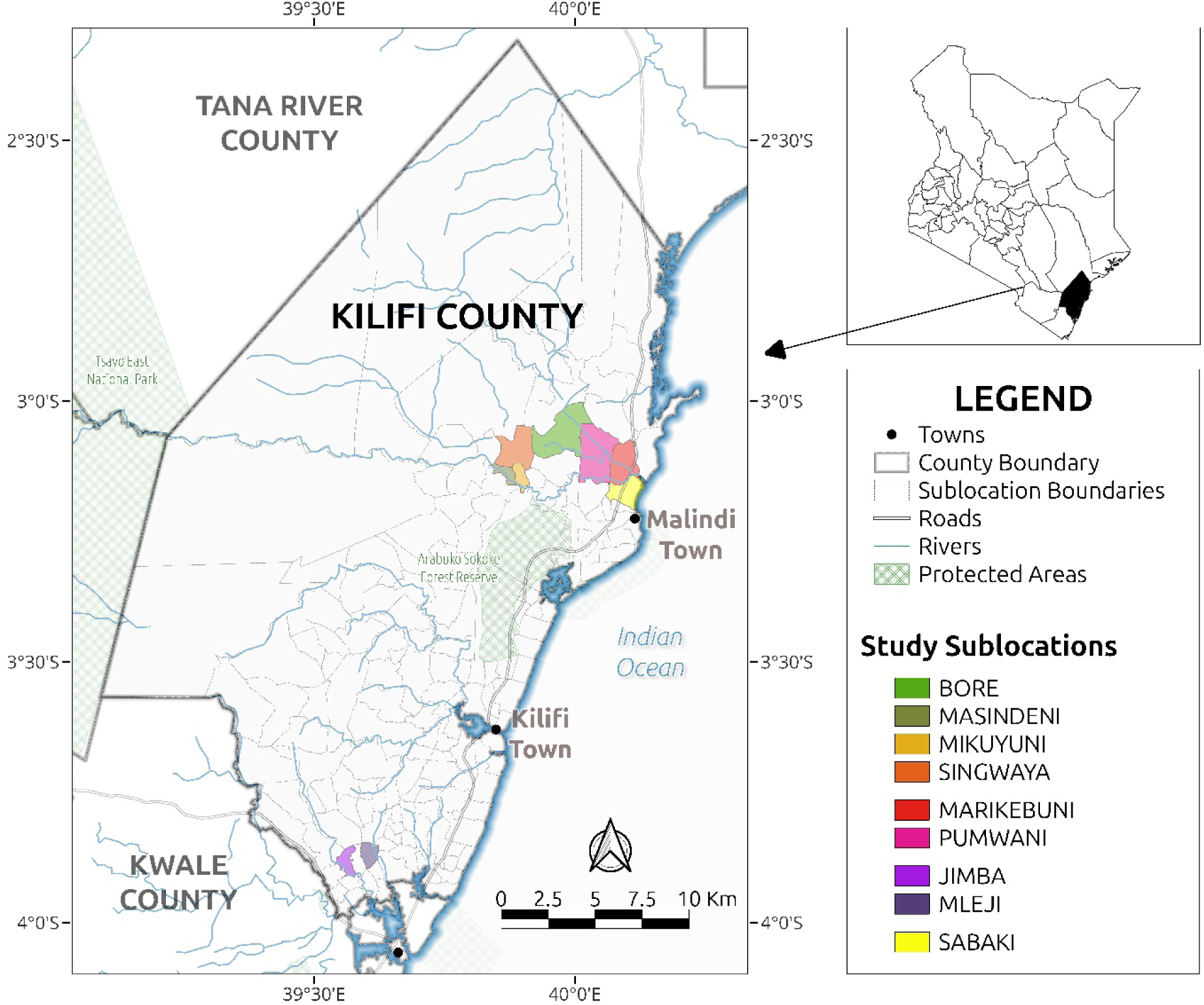
Study area in Kilifi County, Kenya. The map displays Rabai and Magarini sub-counties within Kilifi County, including participating wards and sub-locations. Major administrative boundaries, transport networks, and river systems are shown to contextualise the geographic setting of the study.

### Phase II: FGS assessment

From the 336 women enrolled in Phase I, a stratified random sample of 320 participants was selected using sub-county as the stratification variable to ensure proportional geographic representation. Selection into Phase II was independent of Phase I urine microscopy findings; participants were randomly sampled for FGS assessment regardless of urinary schistosomiasis status, thereby including women with both urine-positive and urine-negative results. Selected participants attended designated village dispensaries within their respective sub-counties for genital specimen collection. A self-sampling approach was employed to enhance privacy and acceptability. Trained female healthcare workers and the principal investigator provided standardised verbal instructions and illustrated guidance. Participants collected genital specimens, including high-vaginal swabs, vaginal swabs, and vaginal secretions, in private rooms at the dispensaries. Of the 320 selected participants, 121 were from Rabai sub-county and 199 from Magarini sub-county. Sixteen women did not proceed to Phase II due to sociocultural concerns related to gynaecological procedures, stigma surrounding reproductive health examinations, or logistical challenges in accessing the dispensaries.

### Final analytic sample

A total of 320 genital swabs were collected. Of these, 291 samples were successfully analysed. However, complete datasets (urine microscopy results, FGS PCR results, and completed questionnaires) were available for 261 participants. Therefore, the final analysis was restricted to these 261 women. Pregnant women and those who had not abstained from sexual intercourse for at least 48 hours before the collection of genital samples were excluded from Phase II participation. The abstinence requirement was implemented to reduce potential contamination or interference with subsequent molecular analyses performed on the genital swabs and was not specific to FGS detection.

### Data Collection Procedures

A structured questionnaire was used to collect sociodemographic and behavioural information, including economic activities, water contact behaviours, education level, and other potential risk factors for *S. haematobium* infection. To evaluate urinary schistosomiasis, participants provided urine samples collected over three consecutive days, which improved diagnostic sensitivity. The urine results from the first phase were included in this analysis to contextualise FGS outcomes. In the second phase, genital samples were gathered at satellite dispensaries. Participants provided high vaginal swabs (HVS) and vaginal swabs through a self-sampling method, ensuring privacy and encouraging higher participation rates. This combined approach of urine and genital sampling enhanced diagnostic accuracy by capturing both urinary and genital forms of schistosomiasis, offering a more complete assessment of the disease burden among women of reproductive age in Kilifi County.

### Urine Collection and Microscopy

Urine collection and microscopy were performed during the first phase of the study. Briefly, urine was collected for three consecutive days. Each participant was requested to give a 20 - 50 ml urine sample between 10 am and 2 pm in a sterile plastic container for three consecutive days. Some of the urine (40ml) collected on the first day was used for the pregnancy determining test and urinalysis. On each of the three days, 10 ml of the urine sample from each participant was filtered through a 12 μm-pore polycarbonate filter and examined for *S. haematobium* eggs under a light microscope (37). The parasite eggs were counted, and the mean egg count was calculated by averaging the total count across the three days. Infection intensity was categorised based on the mean egg count as follows: negative if no eggs were found, low intensity (1 – 49 eggs/ 10 ml urine) and high intensity ( ≥50 eggs/10 ml urine) according to WHO guidelines (38,39) The remaining 40 ml of urine was stored in cryovials for future analysis and transported to the Department of Medical Microbiology and Immunology, University of Nairobi.

### High Vaginal Swab Collection

Each participant was briefly trained on how to self-collect a high vaginal and vaginal swabs. Each participant collected a vaginal swab by holding and inserting a 6-inch sterile vaginal swab in their vagina until the fingers touched the labia minora. The swab was rotated against the vaginal walls for a minimum of 15 repetitions. It was then removed and put back in the swab holder. For the cervical/ high vaginal swab, the participant inserted a sterile 6-inch flocked swab in her vagina until they met noticeable resistance. They then rotated the swab against the vaginal walls 15 times. The swab was withdrawn, the shaft broken, and the swab was put in a sterile screw cap microtube containing 1ml phosphate buffer saline (PBS) and stored at −20°C at the Kilifi County Hospital laboratory. The stored samples were subsequently transported in liquid nitrogen to the Department of Medical Microbiology at the University of Nairobi’s Faculty of Health Sciences for further processing.

### Isolation and PCR Detection of *Schistosoma haematobium* DNA

The QIAamp® DNA Mini Kit was used to extract DNA from high vaginal swabs resuspended in 1 ml of phosphate-buffered saline (QIAGEN, Germany). Proteinase K (20 μl) was added to a microcentrifuge tube to which 200 μl of the sample was applied. The sample was lysed using 200 μl of the AL lysis buffer and then incubated at 56°C for 10 minutes. Binding conditions were adjusted with 200 μl of absolute ethanol. The mixture was transferred to a QIAamp Mini spin column and centrifuged at 8,000 rpm for 1 minute, and the flow-through was discarded. The DNA was washed twice with 500 μl of buffers AW1 and AW2, centrifuged, and the filtrate discarded. To eliminate carryover of buffer AW2, the mini spin columns were dry spun at 13,500 rpm for 1 minute. DNA was eluted with 60 μl of buffer AE. The SensiFAST™ Probe Lo-ROX One-Step Kit was used to perform real-time PCR (Meridian Bioscience, Ohio, USA). The PCR reaction volume (20 μl) consisted of 16 μl master mix and 4 μl DNA. *S. haematobium* DNA was detected through the amplification of the *Dra1* gene sequence, Sh-FW: 5′-GATCTCACCTATCAGACGAAAC-3′; Sh-RV:

5′-TCACAACGATACGACCAAC-3′; Sh-probe:

5’ FAMTGTTGGTGGAAGTGCCTGTTTCGCAA-TAMRA-3’. Amplification results were assessed using cycle threshold (Ct) values produced by the real-time PCR platform. Samples showing detectable amplification within 40 PCR cycles were classified as positive for *S. haematobium* DNA, provided that the positive and negative controls performed as expected. A Ct threshold of 40 cycles was set to ensure high analytical sensitivity and to allow detection of low-intensity infections, which are common in endemic community settings and may be missed by conventional parasitological methods, such as urine microscopy.

Since no universally standardised Ct cut-off exists for *Dra1*-based real-time PCR assays, the positivity threshold was determined based on the assay conditions used in this study. Late amplification signals (Ct ≥35) were interpreted cautiously and considered valid only when amplification curves showed the expected sigmoidal profile and the assay controls were satisfactory.

### Data management and statistical analysis

All data were entered into Microsoft Excel (Microsoft® Corp., Redmond, WA, USA) and subsequently cleaned and validated for completeness and internal consistency. Analyses focused on participants who had complete urine microscopy results, genital PCR results, and questionnaire data (complete-case analysis). The primary outcome was female genital schistosomiasis (FGS), defined as the detection of the *S. haematobium* Dra1 gene sequence by real-time PCR in high vaginal swabs. Prevalence was calculated as the proportion of participants with PCR-confirmed FGS among those included in the final analytical sample. Descriptive statistics summarised sociodemographic, behavioural, and clinical characteristics. Categorical variables were compared using Pearson’s chi-square or Fisher’s exact tests, as appropriate. Continuous variables were assessed using the Wilcoxon rank-sum test. The association between age and mean urinary egg count was evaluated using Spearman’s rank correlation.

### Regression Modelling

A stepwise modelling approach was employed to identify factors associated with FGS. In the initial univariate analyses, each independent variable was assessed for its association with FGS status. Variables with p < 0.25 were considered candidates for multivariable analysis to minimise the risk of excluding potentially relevant predictors. Crude odds ratios (cORs) and 95% confidence intervals (CIs) were estimated using binary logistic regression. Variables meeting the inclusion threshold were entered into a multivariable logistic regression model to estimate adjusted odds ratios (aORs) with 95% CIs. This model aimed to identify independent predictors of FGS while accounting for potential confounding factors. Model diagnostics included the assessment of multicollinearity and overall model fit. No evidence of multicollinearity was observed. Statistical significance was defined as p < 0.05. Binary and multivariable regression analyses were conducted using IBM SPSS Statistics for Windows, Version 26 (IBM Corp., Armonk, NY, USA). Additional analyses, including data visualisation and correlation modelling, were performed using R software (version 4.4.1; R Foundation for Statistical Computing, Vienna, Austria, https://www.r-project.org/>).

### Additional Analyses

To explore the relationship between genital parasite DNA burden and urinary infection intensity, a second-degree polynomial regression model was fitted, with PCR cycle threshold (Ct) values as the dependent variable and mean urinary egg count as the predictor. Differences in Ct values across infection intensity categories (negative, low, high) were evaluated using the Kruskal–Wallis test. Multinomial logistic regression was employed to assess the association between continuous Ct values and infection intensity classification.

### Ethical approval and considerations

The study was conducted in accordance with the Declaration of Helsinki. Ethical approval was obtained from the Kenyatta National Hospital–University of Nairobi Ethics and Research Committee (KNH-ERC) (approval number P334/05/2018) and the National Commission for Science, Technology and Innovation (NACOSTI) (reference number NACOSTI/P/22/1769). Additional administrative permission to conduct the study was granted by the Kilifi County Health Research Committee and the Kilifi County Government through the offices of the County Commissioner and the County Education Office. The study was implemented under the approved protocol titled *Female Genital Schistosomiasis and HIV Infection: The Effect of Schistosoma haematobium on α4β7 Expression among Kenyan Women*. Written informed consent was obtained from all participants before enrolment. Participants aged 15 to 17 years who met the criteria for emancipated minors under Kenyan ethical guidelines provided written informed consent independently, as approved by the KNH-ERC; therefore, parental or guardian consent was not required. All participants were informed of the study’s objectives, procedures, potential risks, and benefits before participation. Confidentiality and anonymity were ensured by using unique study identifiers. Participants who tested positive for *S. haematobium* infection by urine microscopy were treated with praziquantel (Bayer AG, Germany) at a dose of 40 mg/kg, in accordance with Kenyan national treatment guidelines.

## RESULTS

### Sociodemographic characteristics of the study population

Of the 261 participants, 57% were from Magarini sub-county and 43% from Rabai. Participants were distributed across six sub-locations, with the highest representation being in Jimba (27%) in Rabai sub-county and Pumwani (25%) in Magarini sub-county. Most participants were farmers (62%), and 93% had primary education (**Table 1**). The main water source was the river (46%), and 92% reported contact with stagnant water, mainly while washing or fetching water (63%). Most participants reported contacting stagnant water more than twice a week (66%) (**Table 2**). A few participants reported having a history of bilharzia (17%), but only 15% reported receiving treatment. The majority of participants were aged between 36 - 45 years (36%). Reported symptoms included dysuria (28%), abnormal vaginal discharge (26%), bleeding outside menses (13.8%), and bleeding after intercourse (9.6%). Treatment for urinary tract infections (UTIs) was reported by 13%, with 8.8% experiencing recurrent UTIs (**Table 3**).

**Table 1.** Socio-demographic characteristics of study participants (N = 261)

| **Characteristic** | **Category** | **n (%)** |
| --- | --- | --- |
| **Sub-county** | Magarini | 150 (57%) |
|  | Rabai | 111 (43%) |
| **Sub-location** | Marikebuni | 35 (13.4%) |
|  | Garashi | 21 (8.0%) |
|  | Jimba | 71 (27.2%) |
|  | Mleji | 40 (15.3%) |
|  | Pumwani | 64 (24.5%) |
|  | Sabaki | 30 (11.5%) |
| **Age group (years)** | 15–25 | 51 (20%) |
|  | 26–35 | 89 (34%) |
|  | 36–45 | 94 (36%) |
|  | 46–55 | 27 (10%) |
| **Occupation** | Business | 45 (17%) |
|  | Casual labourer | 10 (3.8%) |
|  | Farmer | 161 (62%) |
|  | Housewife | 45 (17%) |
| **Education level** | Primary | 243 (93%) |
|  | Secondary | 18 (6.9%) |

**Table 2.** Water sources and exposure patterns to stagnant water among study participants (N = 261)

| **Characteristic** | **Category** | **n (%)** |
| --- | --- | --- |
| **Source of water for domestic use** | Piped water | 35 (13%) |
|  | Piped water and pond | 4 (1.5%) |
|  | Piped water and river | 22 (8.4%) |
|  | Piped water and well | 1 (0.4%) |
|  | Pond | 52 (20%) |
|  | River | 121 (46%) |
|  | River and pond | 2 (0.8%) |
|  | Well | 20 (7.7%) |
|  | Well and river | 4 (1.5%) |
| **Contact with stagnant water** | Yes | 241 (92%) |
|  | No | 20 (8%) |
| **Mode of contact** | None | 20 (7.7%) |
|  | When bathing | 31 (12%) |
|  | When walking | 17 (6.5%) |
|  | When washing/fetching water | 165 (63%) |
|  | Washing/fetching water<br>and bathing | 24 (9.2%) |
|  | Washing/fetching water<br>and walking | 4 (1.5%) |
| **Frequency of contact** | Less than twice a week | 18 (6.9%) |
|  | More than twice a week | 172 (66%) |
|  | Seasonally | 51 (20%) |
|  | None | 20 (7.7%) |

**Table 3.** History of schistosomiasis, treatment, and gynaecological and urinary symptoms among study participants (N = 261)

| **Characteristic** | **Category** | **n (%)** |
| --- | --- | --- |
| **History of schistosomiasis** | Yes | 45 (17%) |
|  | No | 216 (83%) |
| **Received schistosomiasis treatment** | Yes | 38 (15%) |
|  | No | 223 (85%) |
| **Bleeding outside menses (last 6 months)** | None | 225 (86.2%) |
|  | More than 6 months ago | 13 (5.0%) |
|  | Within the last 6 months | 12 (4.6%) |
|  | Presently | 11 (4.2%) |
| **Dysuria (last 3 months)** | Yes | 72 (28%) |
|  | No | 189 (72%) |
| **Bleeding after intercourse** | Yes | 25 (9.6%) |
|  | No | 236 (90.4%) |
| **Abnormal vaginal discharge (last 3 months)** | Yes | 67 (26%) |
|  | No | 194 (74%) |
| **Treated for UTI (last 3 months)** | Yes | 34 (13%) |
|  | No | 227 (87%) |
| **UTI recurrence** | Yes | 23 (8.8%) |
|  | No | 238 (90.2%) |

### Prevalence of female genital schistosomiasis in Kilifi County

Overall, the prevalence of FGS was 36.0% (94/261) in Magarini and Rabai sub-counties. In Magarini, 150 women were enrolled, whereas 111 women were recruited from Rabai. Between the two sub-counties, Magarini had the highest prevalence of FGS, standing at 43.3% (65/150), whereas Rabai had a FGS prevalence of 26.1% (29/111) (**Table 4**, **Figure 2a)**. Amongst the wards, Sabaki had the highest prevalence at 56.7% (17/30), followed by Magarini 44.4% (44/99), Ruruma 26.1% (29/111) and Garashi 19.0% (4/21) (**Figure 2c**). Out of all the sub-locations, Marikebuni recorded the highest prevalence at 68.6% (24/35), followed by Pumwani at 31.3% (20/64), Jimba at 26.8% (19/71) and Mleji at 25.0% (10/40) (**Table 4**).

**Figure 2:**
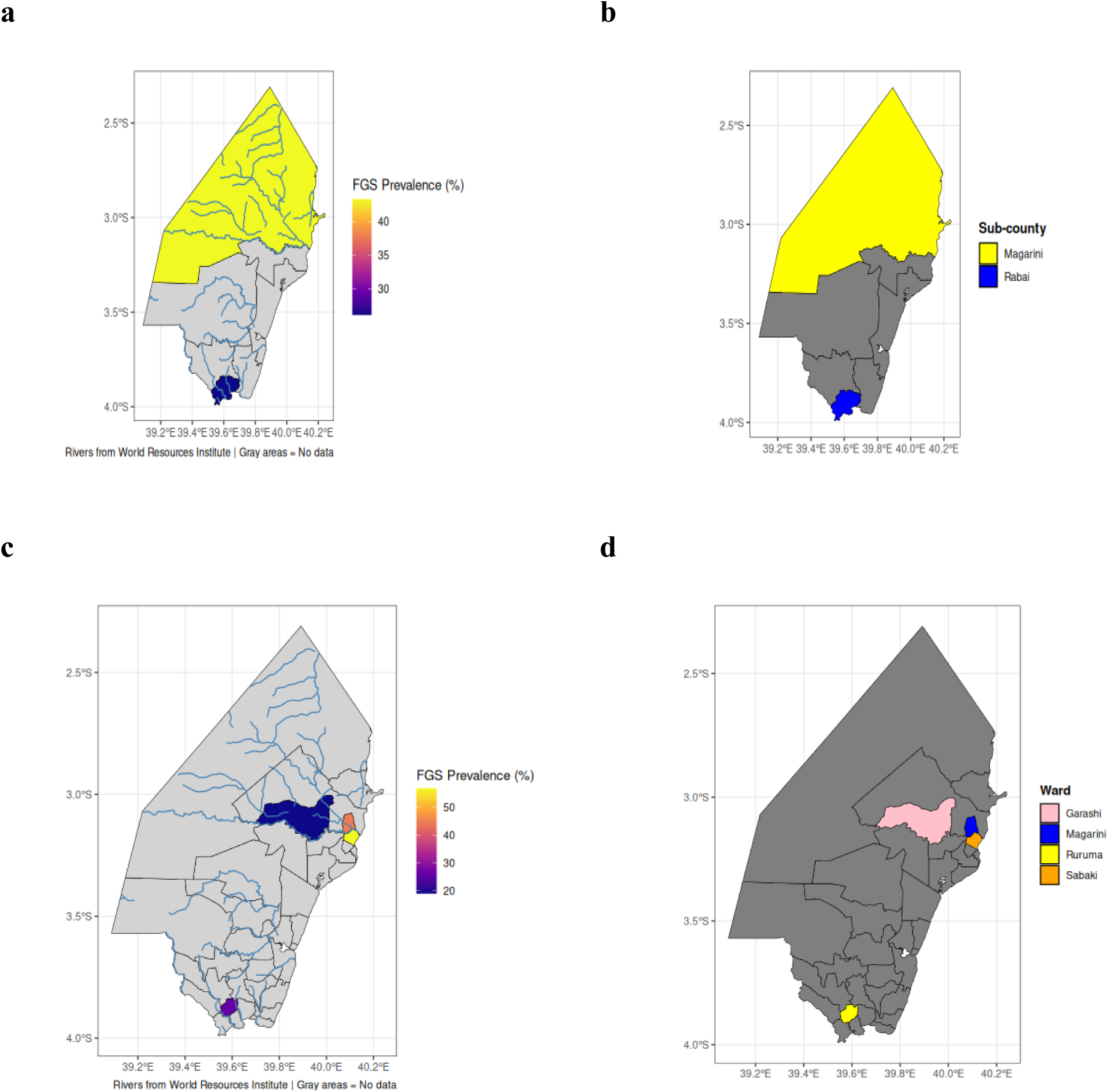
Geospatial distribution of female genital schistosomiasis in selected areas of Kilifi County, Kenya (40,41): (a) Sub-county level, (b) Sub-county reference map, (c) Ward level, (d) Ward reference map.

**Table 4:** Prevalence of female genital schistosomiasis among women of reproductive age in Kilifi County.

| Characteristic | Negative, N = 167 | Positive, N = 94 | Total, N = 261<sup>1</sup> |
| --- | --- | --- | --- |
| **Sub-county** |  |  |  |
| Magarini | 85 | 65 | 150 (43.3%) |
| Rabai | 82 | 29 | 111 (26.1%) |
| **Ward** |  |  |  |
| Magarini | 55 | 44 | 99 (44.4%) |
| Ruruma | 82 | 29 | 111 (26.1%) |
| Garashi | 17 | 4 | 21 (19.0%) |
| Sabaki | 13 | 17 | 30 (56.7%) |
| **Sub-location** |  |  |  |
| Marikebuni | 11 | 24 | 35 (68.6%) |
| Jimba | 52 | 19 | 71 (26.8%) |
| Mleji | 30 | 10 | 40 (25%) |
| Pumwani | 44 | 20 | 64 (31.3%) |
| *l* n (%) |  |  |  |

### Urinalysis parameters associated with FGS

Haematuria was significantly correlated with FGS (p<0.05) (**Table 5**), and women in Magarini were more likely to present with it in comparison to those in Rabai (p<0.05) (**Table 6**). Additionally, the presence of *S. haematobium* eggs in urine was associated with FGS, with eggs being observed in 13% (34/261) of the participants, including a few women who didn’t have FGS (p<0.001). The prevalence of urogenital schistosomiasis and FGS co-occurrence was 28% (26/94) (**Table 5**).

**Table 5:** Urinalysis results in relation to female genital schistosomiasis status among women of reproductive age in Kilifi County.

| **Characteristic** | Negative N=167<sup>1</sup> | Positive N=94<sup>1</sup> | p-value<sup>2</sup> |
| --- | --- | --- | --- |
| Haematuria |  |  | 0.031 |
| **Negative** | 163 (98%) | 86 (91.5%) |  |
| **Positive** | 4 (2.4%) | 8 (8.5%) |  |
| Nitrites |  |  | >0.9 |
| **Negative** | 161 (96%) | 91 (97%) |  |
| **Positive** | 6 (3.6%) | 3 (3.2%) |  |
| Leucocytes |  |  | 0.13 |
| **2+** | 5 (3.0%) | 4 (4.3%) |  |
| **3+** | 10 (6.0%) | 12 (13%) |  |
| **Negative** | 152 (91%) | 78 (83%) |  |
| Presence of *S. haematobium* eggs in urine |  |  | <0.001 |
| **Negative** | 159 (95%) | 68 (72%) |  |
| **Positive** | 8 (4.8%) | 26 (28%) |  |
| <sup>1</sup>n (%) | <sup>2</sup>Pearson's Chi-squared test; Fisher's exact test |  |  |

**Table 6:**
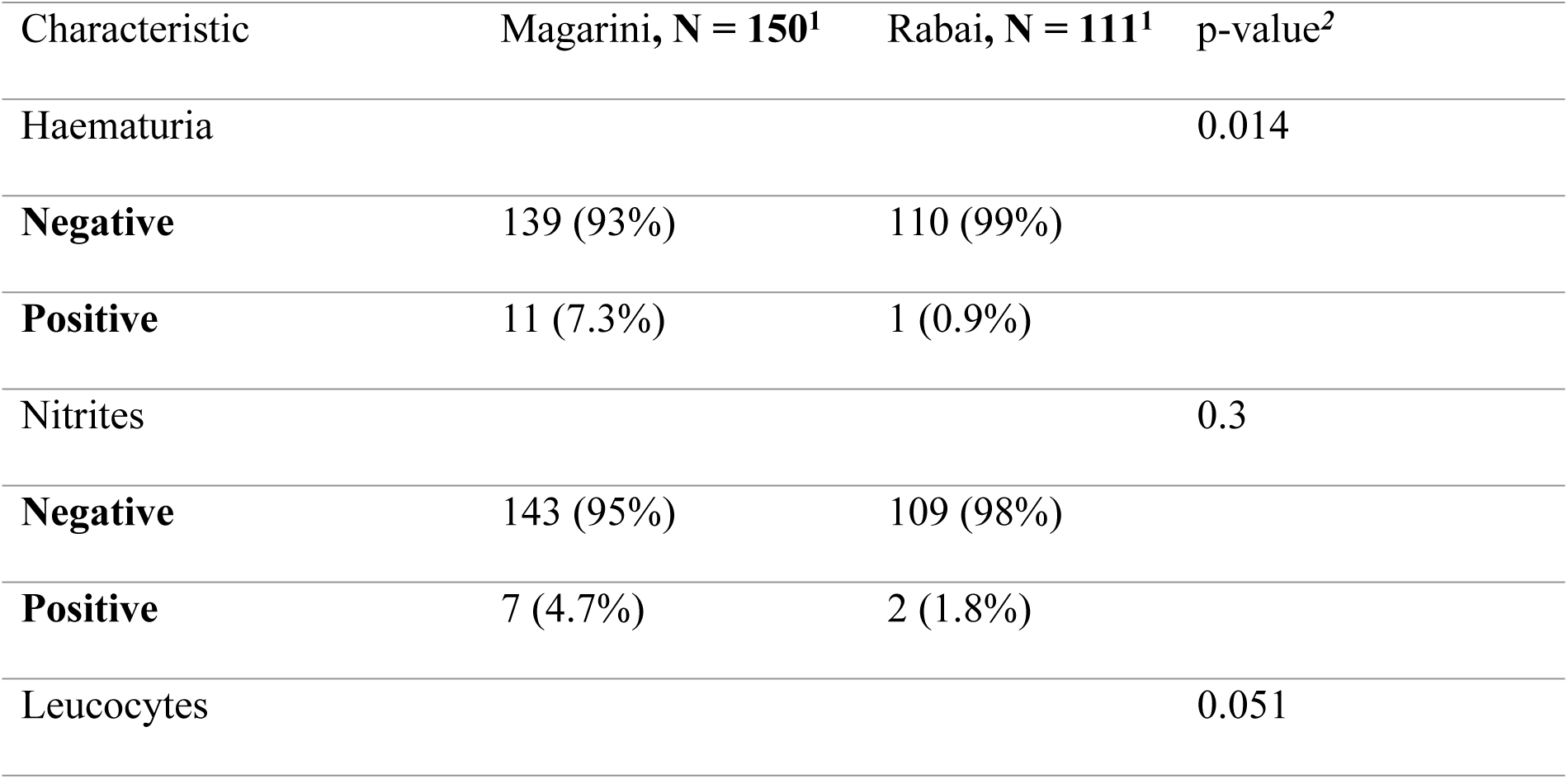
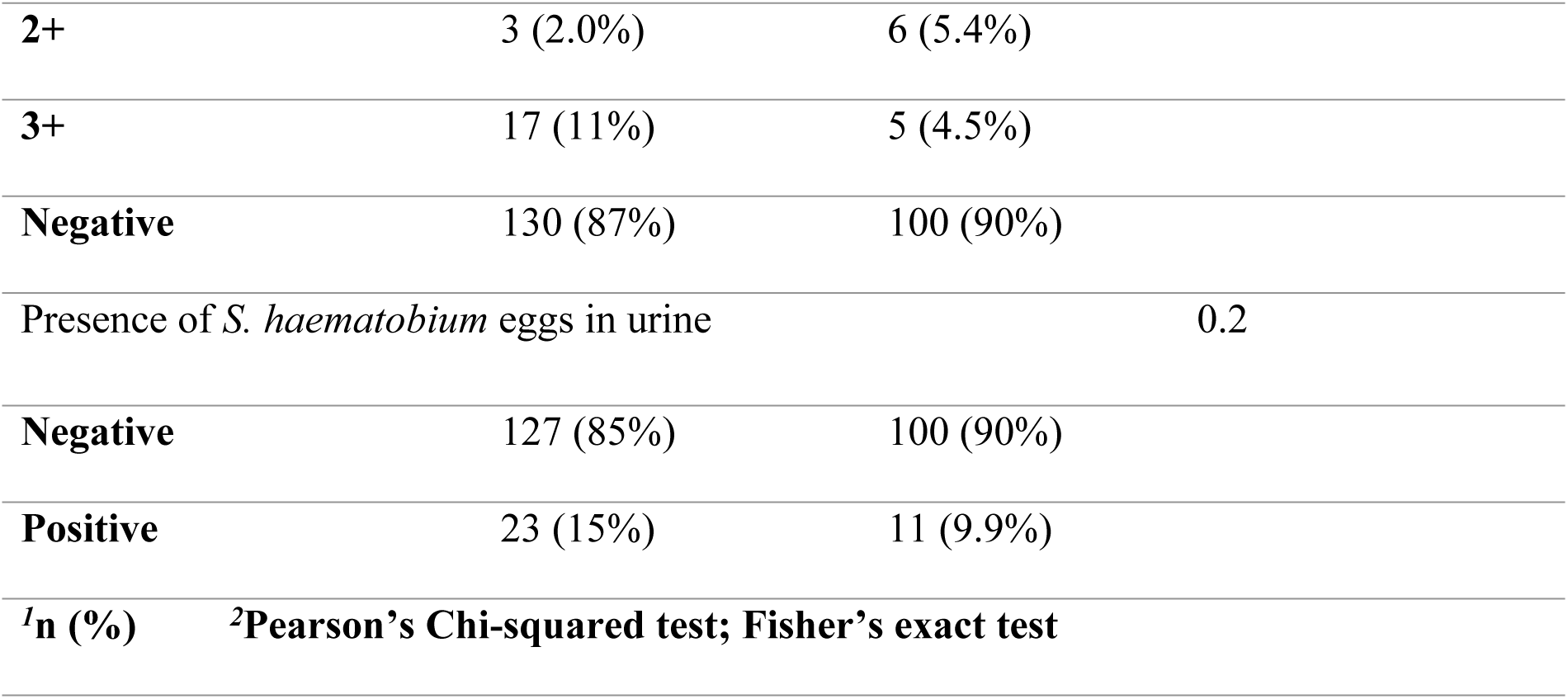
Urinalysis results in Magarini and Rabai sub-counties.

### FGS can occur without pathophysiological involvement in the urinary tract

*S. haematobium* eggs were not detected in urine samples from 72% of the women who were positive by PCR (**Table 5**). In Rabai sub-county, 76% (22/29) of the women who had FGS tested negative for *S. haematobium* eggs, whereas in Magarini sub-county, 71% (46/65) of the women had no eggs in urine. The same pattern was observed in the wards as follows: 94% (16/17) in Sabaki, 90% (9/10) in Mleji, 68% (13/19) in Jimba, 67% (16/24) in Marikebuni, 60% (12/20) in Pumwani and 50% (2/4) in Garashi.

### PCR is sensitive in detecting FGS and *S. haematobium* infection intensity in comparison to urine microscopy

PCR-based detection identified a substantially higher prevalence of FGS than urine microscopy. The prevalence of FGS detected by PCR was 36.0%, whereas urinary egg excretion detected by microscopy was 13.0%. Overall concordance between the two diagnostic methods was 70.9% (185/261), comprising 26 concordant positive and 159 concordant negative cases. Among women with PCR-confirmed FGS, 72% had no detectable *S. haematobium* eggs in urine. A polynomial regression model was used to evaluate the relationship between Ct values and mean urinary egg counts. The first-degree polynomial term was not statistically significant (p = 0.052), whereas the second-degree term was significant (p < 0.001), indicating a non-linear relationship between Ct value and mean egg count (Figure 3a). The model explained a modest proportion of the variation (R² = 9.6%, adjusted R² = 8.9%). Ct values differed significantly across infection intensity categories (Chi-squared = 13.969, p = 0.001). Multinomial logistic regression analysis revealed a significant association between Ct value and infection intensity categories. Each unit increase in Ct value was linked to higher odds of being classified into both low infection intensity (OR = 1.039, p = 0.0018) and high infection intensity categories (OR = 1.049, p = 0.0305) compared to participants with no detectable eggs. Notably, some participants who had no detectable *S. haematobium* eggs in their urine displayed elevated Ct values (>35), suggesting the presence of low levels of parasite DNA despite the absence of detectable egg excretion. In contrast, participants with higher urinary egg counts typically exhibited lower Ct values, reflecting greater parasite DNA loads. Importantly, PCR identified a significant proportion of infections that urine microscopy failed to detect, underscoring the enhanced sensitivity of molecular methods for identifying low-intensity infections that may be overlooked by conventional parasitological techniques (**Figure 3b–c**).

**Figure 3.**
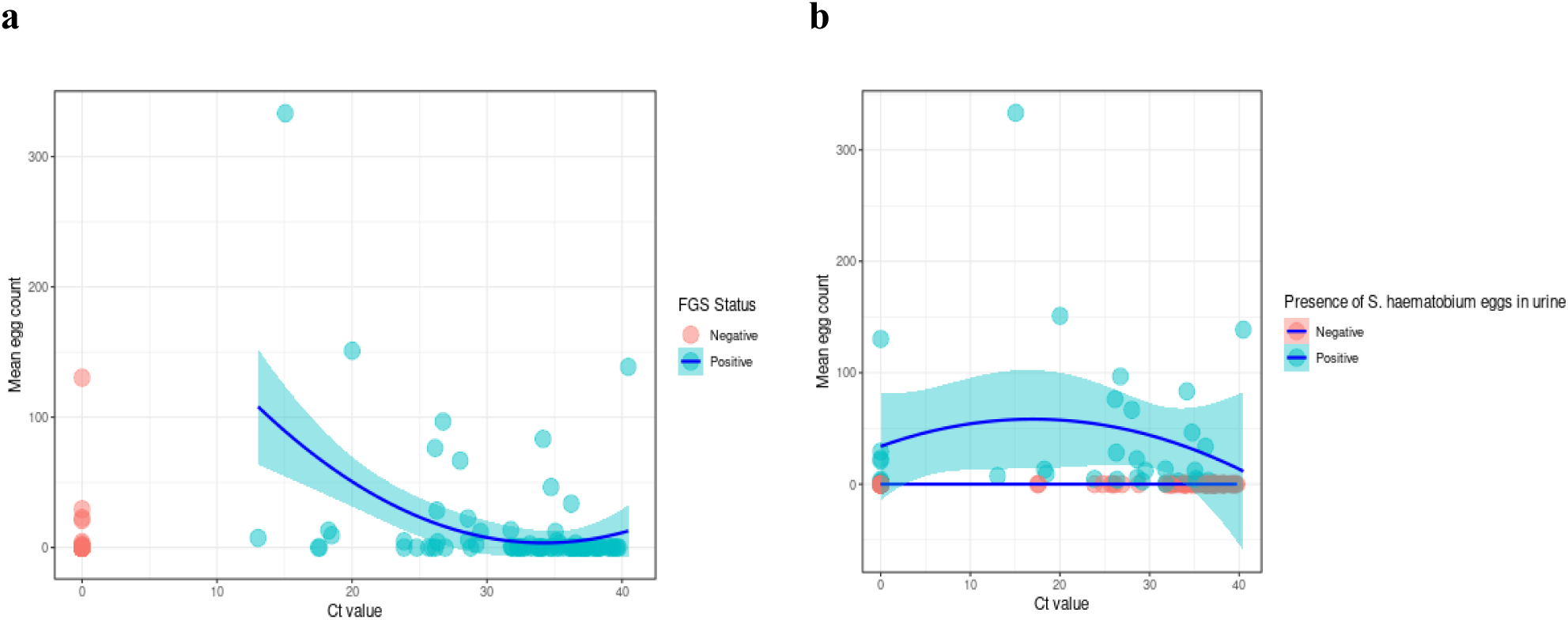
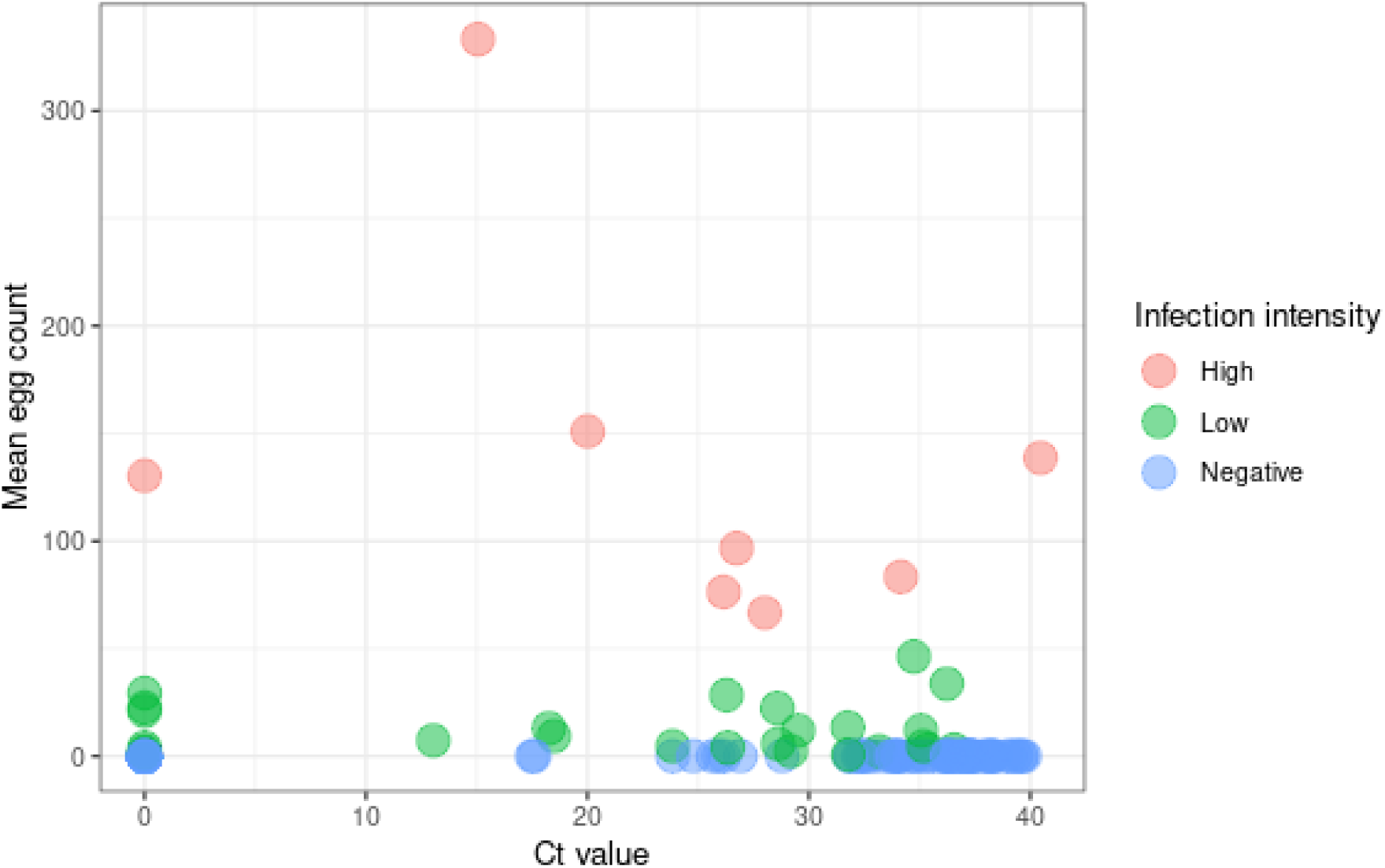
Comparison of PCR and urine microscopy for detection of *Schistosoma haematobium*: (a) Relationship between mean urinary egg count and PCR cycle threshold (Ct) value showing a non-linear polynomial fit with 95% confidence intervals, (b) Distribution of Ct values according to microscopy detection status, and (c) Ct values across urinary infection intensity categories.

### Risk factors associated with FGS among women of reproductive age

#### Sociodemographic factors by infection status

Sociodemographic characteristics stratified by infection status are presented in **Table 7**. Infection status differed significantly by sub-county, with a higher proportion of infected participants residing in Magarini compared with uninfected participants (65/94 [69%] vs 85/167 [51%]), whereas Rabai had a higher proportion of uninfected participants (82/167 [49%] vs 29/94 [31%]; p = 0.004). Significant variation was also observed across sub-locations (p < 0.001), with infected participants more frequently coming from Marikebuni (24/94 [26%] vs 11/167 [6.6%]) and Sabaki (17/94 [18%] vs 13/167 [7.8%]), while uninfected participants were more commonly from Jimba (51/167 [31%] vs 19/94 [20%]) and Pumwani (44/167 [26%] vs 20/94 [21%]).

**Table 7.**
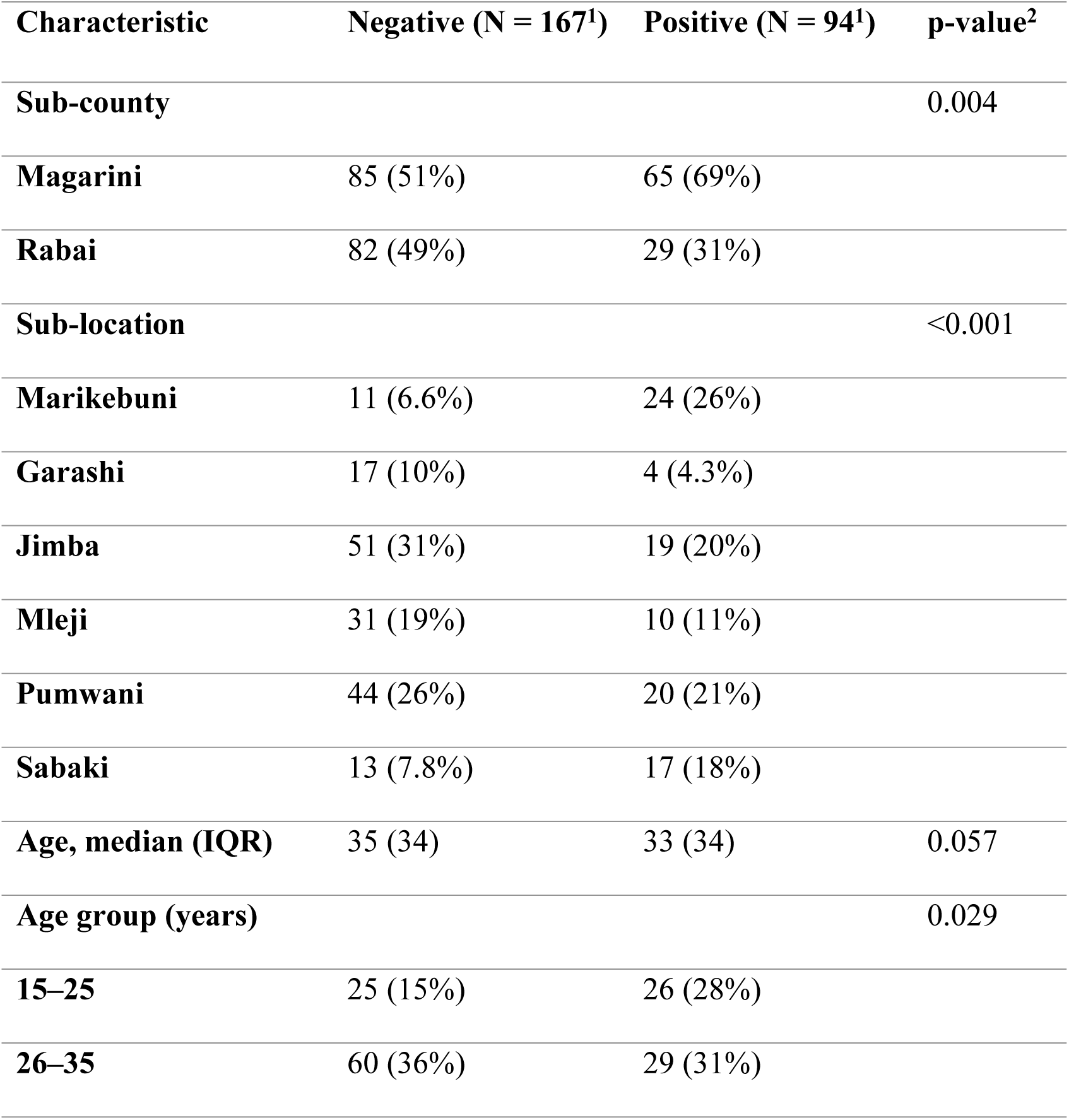
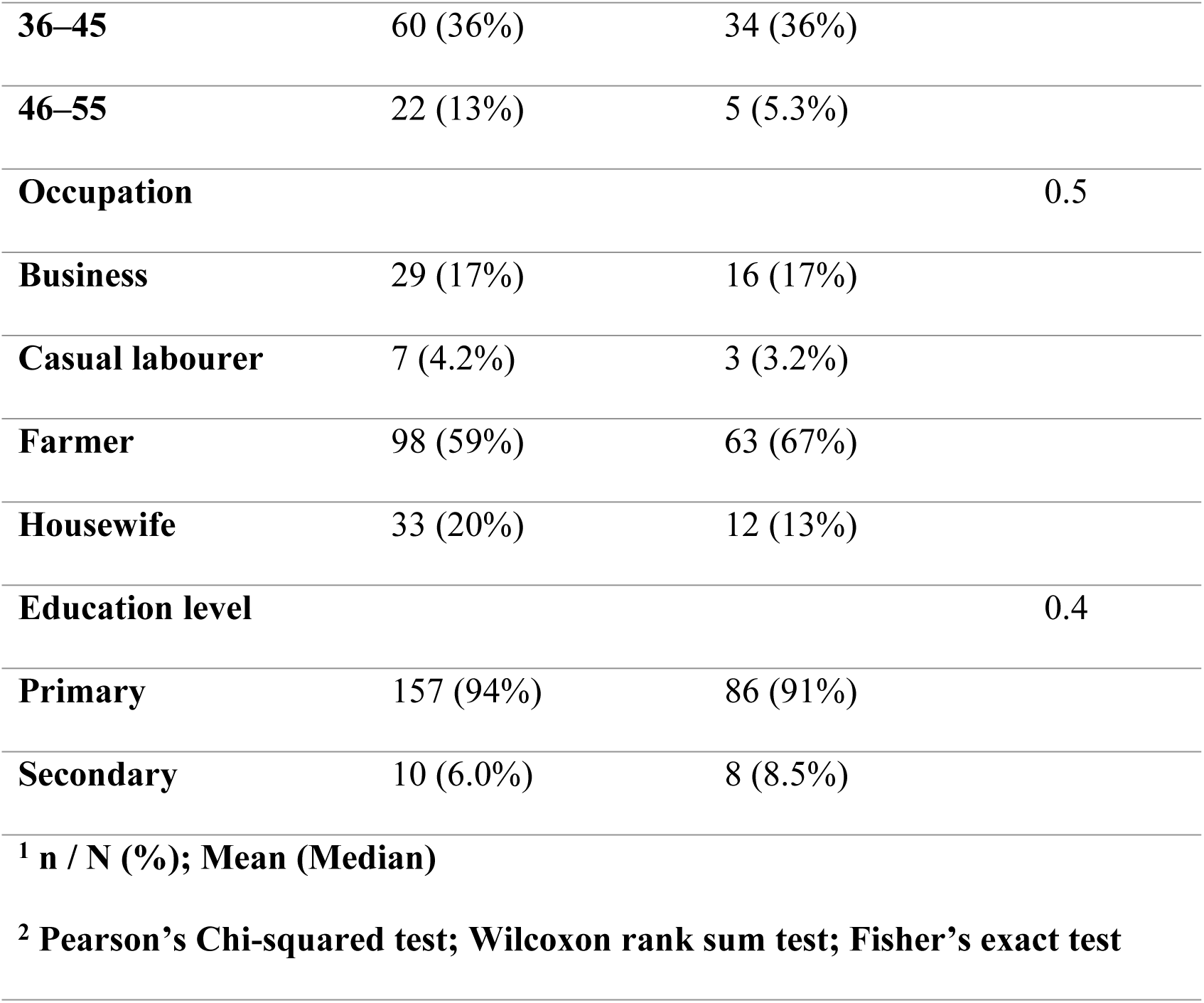
Socio-demographic characteristics by infection status.

Median age was slightly lower among infected participants than uninfected participants (33 [IQR 34] vs 35 [IQR 34]), although this difference did not reach statistical significance (p = 0.057). However, the distribution of age groups differed significantly by infection status (p = 0.029), with women aged 15–25 years representing a higher proportion of infected participants (26/94 [28%] vs 25/167 [15%]), while women aged 46–55 years were less frequently infected (5/94 [5.3%] vs 22/167 [13%]). Occupation and education level did not differ significantly by infection status. Farming was the most commonly reported occupation among both uninfected (98/167 [59%]) and infected participants (63/94 [67%]), and primary education was the predominant level reported in both groups (157/167 [94%] vs 86/94 [91%]).

#### Water sources, behavioural exposures, and contact with stagnant water by infection status

Water sources and exposure to stagnant water stratified by infection status are presented in **Table 8**. River water was the most commonly reported domestic water source in both groups (73/167 [44%] of uninfected participants and 48/94 [51%] of infected participants). The distribution of domestic water sources did not differ significantly by infection status (p = 0.40). Contact with stagnant water was reported by 150/167 (90%) of uninfected participants and 91/94 (97%) of infected participants, representing a statistically significant difference between the groups (p = 0.042). Washing or fetching water was the most frequently reported mode of contact in both groups; however, differences in the mode of contact did not reach statistical significance (p = 0.054). Frequent contact with stagnant water (more than twice per week) was reported by 106/167 (63%) of uninfected participants and 66/94 (70%) of infected participants. The frequency of contact did not differ significantly between the groups (p = 0.20).

**Table 8.**
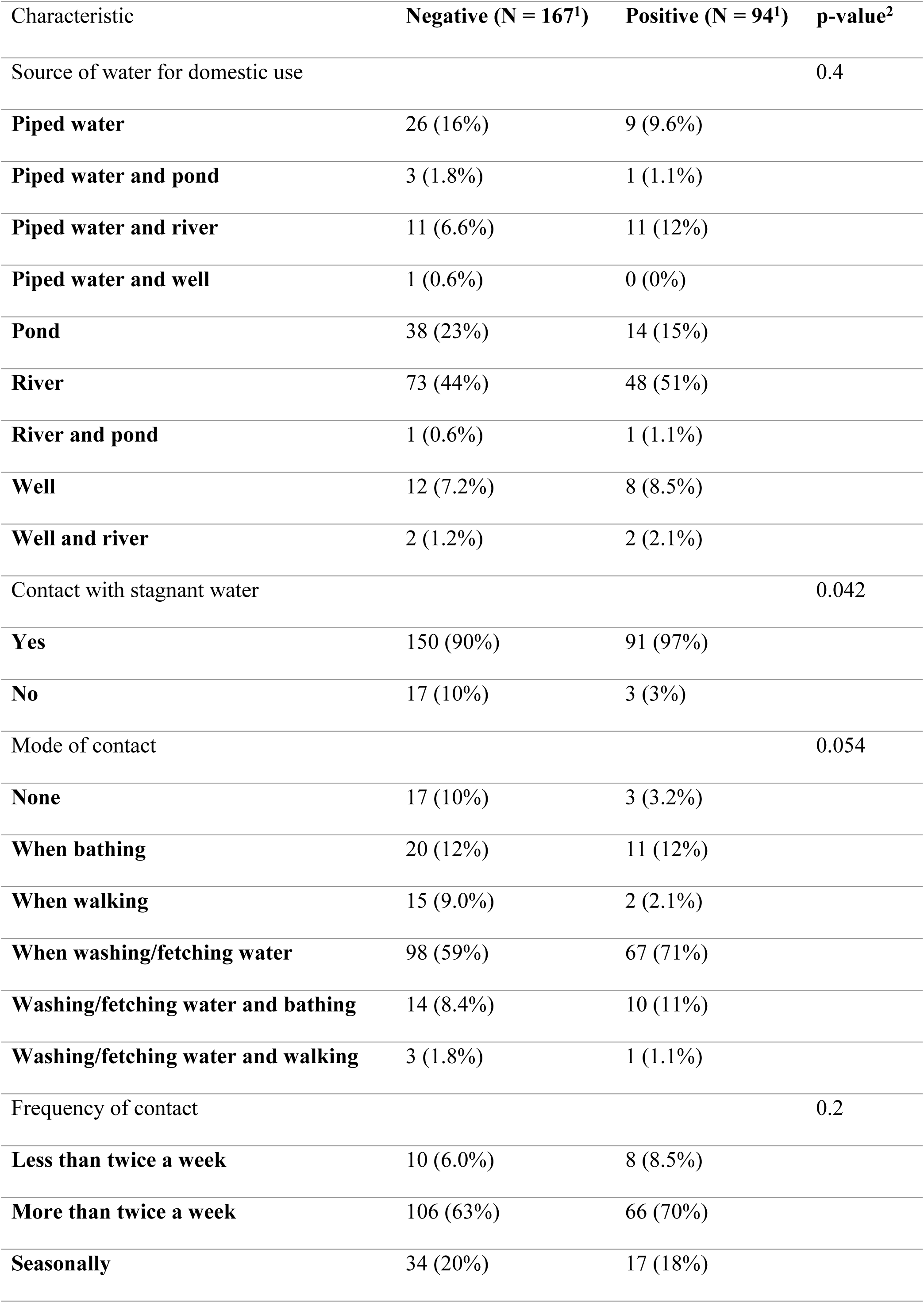
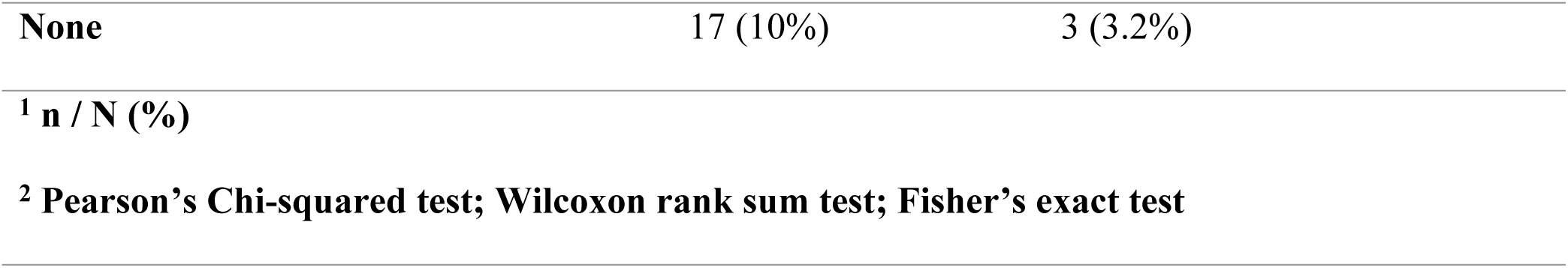
Water sources and exposure to stagnant water by infection status.

| Characteristic | Negative (N = 167<sup>1</sup>) | Positive (N = 94<sup>1</sup>) | p-value<sup>2</sup> |
| --- | --- | --- | --- |
| Source of water for domestic use |  |  | 0.4 |
| **Piped water** | 26 (16%) | 9 (9.6%) |  |
| **Piped water and pond** | 3 (1.8%) | 1 (1.1%) |  |
| **Piped water and river** | 11 (6.6%) | 11 (12%) |  |
| **Piped water and well** | 1 (0.6%) | 0 (0%) |  |
| **Pond** | 38 (23%) | 14 (15%) |  |
| **River** | 73 (44%) | 48 (51%) |  |
| **River and pond** | 1 (0.6%) | 1 (1.1%) |  |
| **Well** | 12 (7.2%) | 8 (8.5%) |  |
| **Well and river** | 2 (1.2%) | 2 (2.1%) |  |
| Contact with stagnant water |  |  | 0.042 |
| **Yes** | 150 (90%) | 91 (97%) |  |
| **No** | 17 (10%) | 3 (3%) |  |
| Mode of contact |  |  | 0.054 |
| **None** | 17 (10%) | 3 (3.2%) |  |
| **When bathing** | 20 (12%) | 11 (12%) |  |
| **When walking** | 15 (9.0%) | 2 (2.1%) |  |
| **When washing/fetching water** | 98 (59%) | 67 (71%) |  |
| **Washing/fetching water and bathing** | 14 (8.4%) | 10 (11%) |  |
| **Washing/fetching water and walking** | 3 (1.8%) | 1 (1.1%) |  |
| Frequency of contact |  |  | 0.2 |
| **Less than twice a week** | 10 (6.0%) | 8 (8.5%) |  |
| **More than twice a week** | 106 (63%) | 66 (70%) |  |
| **Seasonally** | 34 (20%) | 17 (18%) |  |

|  |  |  |
| --- | --- | --- |
| None | 17 (10%) | 3 (3.2%) |
| <sup>1</sup> n / N (%) |  |  |
| <sup>2</sup> Pearson's Chi-squared test; Wilcoxon rank sum test; Fisher's exact test |  |  |

#### Self-reported schistosomiasis-related symptoms, disease and treatment history by infection status

Schistosomiasis-related factors and clinical symptoms stratified by infection status are presented in **Table 9**. A history of bilharzia was more frequently reported among infected participants compared with uninfected participants (25/94 [27%] vs 20/167 [12%]; p = 0.003). Similarly, prior treatment for bilharzia was more common among infected participants (20/94 [21%]) than among uninfected participants (18/167 [11%]; p = 0.021). Marked differences in urinary infection intensity were observed by infection status (p < 0.001). High-intensity infection was detected in 7/94 (7.4%) of infected participants compared with 1/167 (0.6%) of uninfected participants, while low-intensity infection occurred in 19/94 (20%) and 7/167 (4.2%) of participants, respectively. The presence of *S. haematobium* eggs in urine also differed significantly between groups, with eggs detected in 26/94 (28%) of infected participants compared with 8/167 (4.8%) of uninfected participants (p < 0.001). Median egg counts were higher among infected participants than among uninfected participants (12 [IQR 0] vs 1 [IQR 0]; p < 0.001). Bleeding outside of menses and dysuria were more frequently reported among infected participants (p < 0.001 for both), while bleeding after intercourse and abnormal vaginal discharge did not differ significantly between the groups.

**Table 9.**
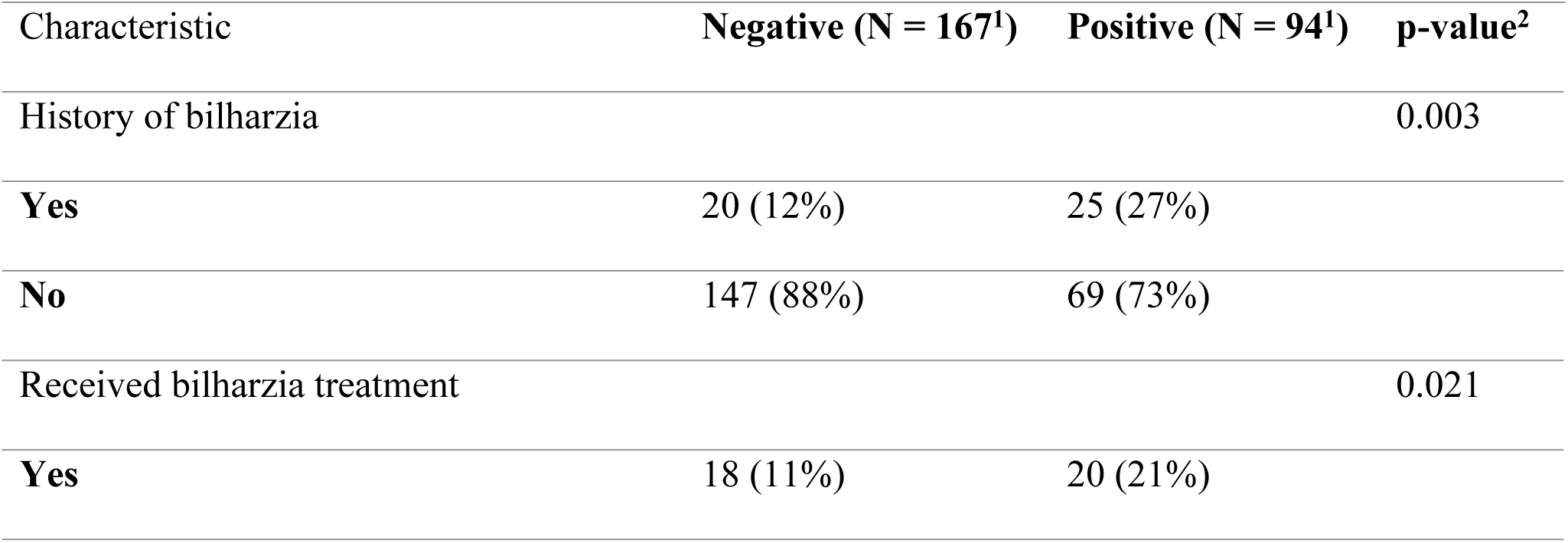
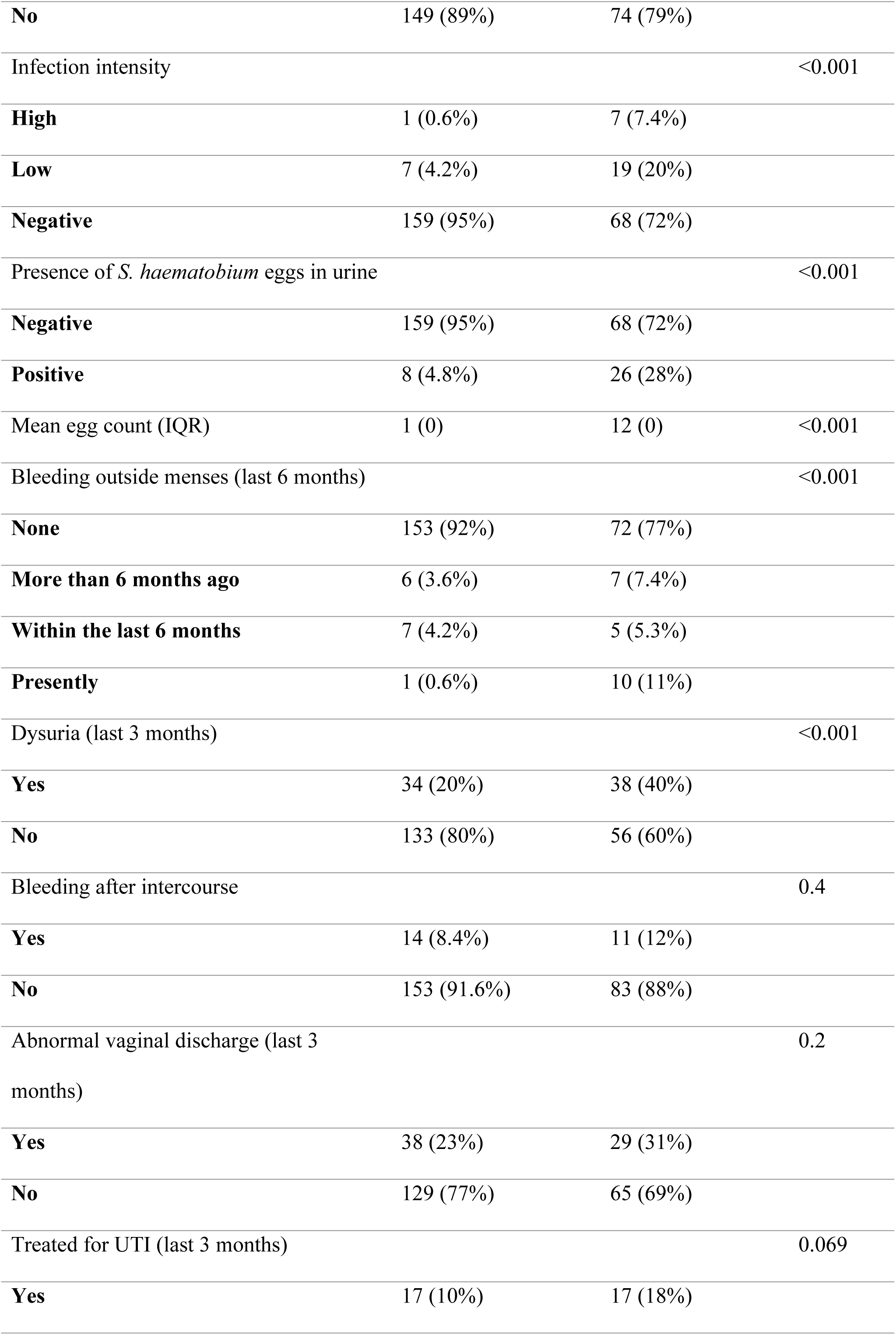
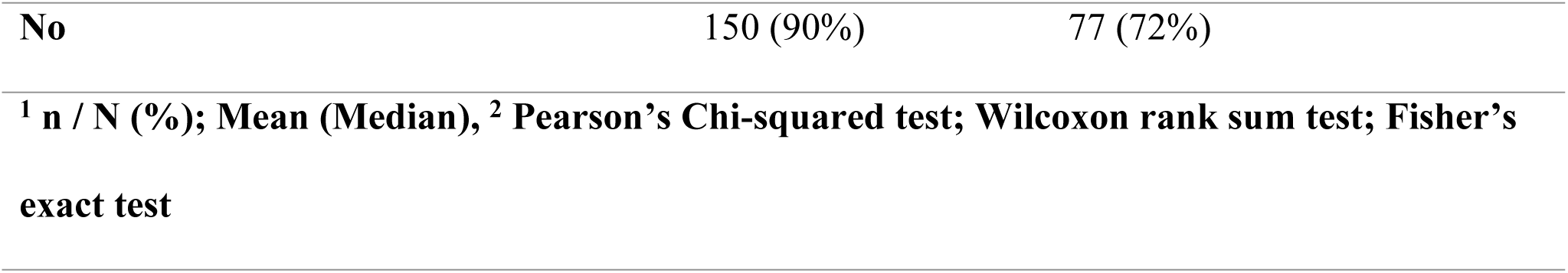
Schistosomiasis-related factors and clinical symptoms by infection status.

| Characteristic | Negative (N = 167<sup>1</sup>) | Positive (N = 94<sup>1</sup>) | p-value<sup>2</sup> |
| --- | --- | --- | --- |
| History of bilharzia |  |  | 0.003 |
| Yes | 20 (12%) | 25 (27%) |  |
| No | 147 (88%) | 69 (73%) |  |
| Received bilharzia treatment |  |  | 0.021 |
| Yes | 18 (11%) | 20 (21%) |  |
| **No** | 149 (89%) | 74 (79%) |  |
| Infection intensity |  |  | <0.001 |
| **High** | 1 (0.6%) | 7 (7.4%) |  |
| **Low** | 7 (4.2%) | 19 (20%) |  |
| **Negative** | 159 (95%) | 68 (72%) |  |
| Presence of *S. haematobium* eggs in urine |  |  | <0.001 |
| **Negative** | 159 (95%) | 68 (72%) |  |
| **Positive** | 8 (4.8%) | 26 (28%) |  |
| Mean egg count (IQR) | 1 (0) | 12 (0) | <0.001 |
| Bleeding outside menses (last 6 months) |  |  | <0.001 |
| **None** | 153 (92%) | 72 (77%) |  |
| **More than 6 months ago** | 6 (3.6%) | 7 (7.4%) |  |
| **Within the last 6 months** | 7 (4.2%) | 5 (5.3%) |  |
| **Presently** | 1 (0.6%) | 10 (11%) |  |
| Dysuria (last 3 months) |  |  | <0.001 |
| **Yes** | 34 (20%) | 38 (40%) |  |
| **No** | 133 (80%) | 56 (60%) |  |
| Bleeding after intercourse |  |  | 0.4 |
| **Yes** | 14 (8.4%) | 11 (12%) |  |
| **No** | 153 (91.6%) | 83 (88%) |  |
| Abnormal vaginal discharge (last 3 months) |  |  | 0.2 |
| **Yes** | 38 (23%) | 29 (31%) |  |
| **No** | 129 (77%) | 65 (69%) |  |
| Treated for UTI (last 3 months) |  |  | 0.069 |
| **Yes** | 17 (10%) | 17 (18%) |  |

|  |  |  |
| --- | --- | --- |
| No | 150 (90%) | 77 (72%) |
| <sup>1</sup> n / N (%); Mean (Median), <sup>2</sup> Pearson's Chi-squared test; Wilcoxon rank sum test; Fisher's exact test |  |  |

#### Differences in sociodemographic and behavioural factors between sub-counties

Differences in sociodemographic and behavioural characteristics between sub-counties are presented in **Table 10**. The mode of contact with stagnant water, frequency of contact with stagnant water, occupation, and source of domestic water differed significantly between Rabai and Magarini sub-counties (p < 0.001 for all). Washing or fetching water was the most commonly reported mode of contact in both sub-counties. Frequent contact with stagnant water was reported more often in Magarini than in Rabai. Farming was the predominant occupation in both sub-counties, but was more prevalent in Magarini. River water was the primary source of domestic water in Magarini, whereas pond water was more commonly reported as the source in Rabai.

**Table 10:**
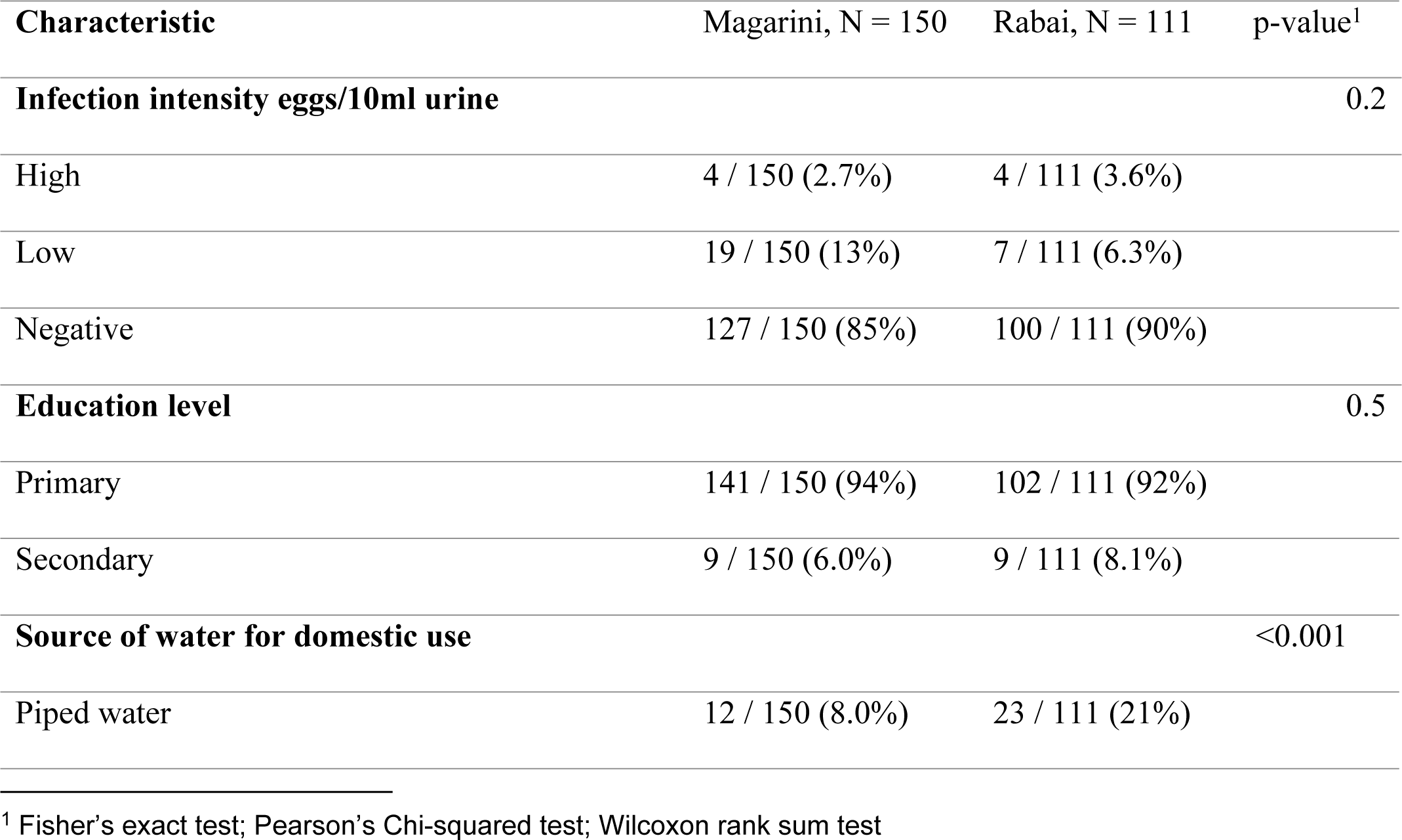
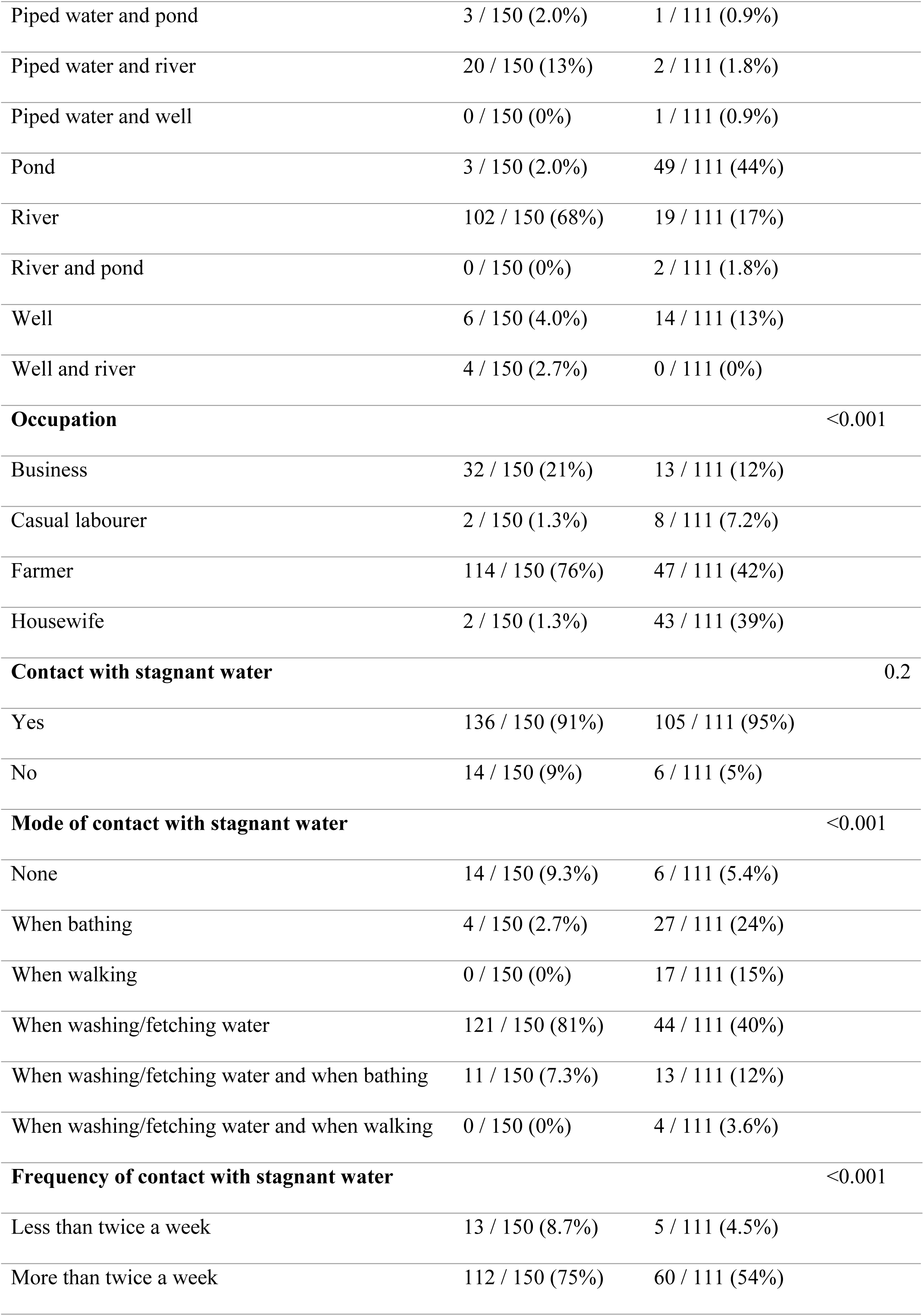
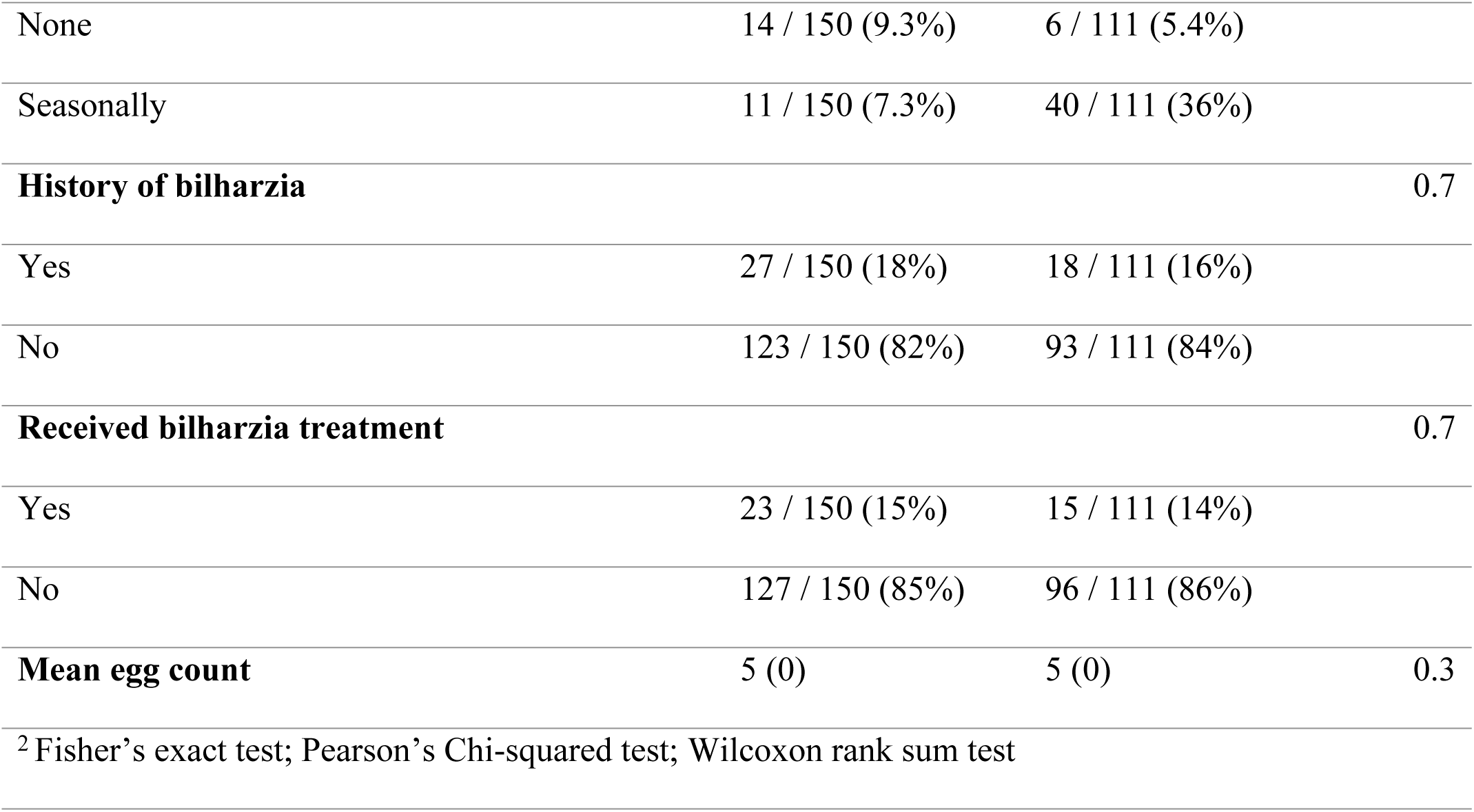
Characteristics associated with female genital schistosomiasis infection status in Magarini and Rabai sub-counties.

### Logistic Binary Regression analysis for factors associated with FGS

In crude logistic regression analysis (**Table 11**), several variables were associated with FGS. These included age group, sub-location of residence, mode of contact with stagnant water, prior treatment for schistosomiasis, bleeding outside of menses, recurrent urinary tract infections, and urinary infection intensity. Notably, light and heavy urinary infection intensities were strongly associated with FGS in the crude analysis. These variables were subsequently considered for inclusion in the multivariable model.

**Table 11:** Binary Logistic Regression Analysis of Independent Predictors of Female Genital Schistosomiasis (FGS) among Study Participants.

| Variable | Total Sample<br>(n = 261) | FGS Positive (n =<br>94 ) | FGS<br>Negative (n<br>= 167 ) | cOR (95% CI) | p-value |
| --- | --- | --- | --- | --- | --- |
| Age -group |  |  |  |  | 0.035 |
| **15 - 25** | 51 (19.5%) | 26 (27.7%) | 25 (15.0%) | 0.219 (0.072 – 0.667) | 0.008 |
| **26 - 35** | 89 (34.1%) | 29 (30.9%) | 60 (35.9%) | 0.470 (0.162 – 1.367) | 0.166 |
| **36 - 45** | 94 (36.0%) | 34 (36.2%) | 60 (35.9%) | 0.401 (0.139 – 1.156) | 0.091 |
| **46 - 55** | 27 (10.3%) | 5 (5.3%) | 22 (13.2%) | **Ref** |  |
| Ward of residence |  |  |  |  | <0.0001 |
| **Jimba** | 71(27.2%) | 19 (20.3%) | 52 (31.1%) | 0.644 (0.192 – 2.158) | 0.476 |
| **Mleji** | 40 (15.3%) | 10 (10.6%) | 30 (18.0%) | 0.706 (0.192 – 2.599) | 0.600 |
| **Pumwani** | 64 (24.5%) | 20 (21.3%) | 44 (26.3%) | 0.518 (0.154 – 1.737) | 0.286 |
| **Marikebuni** | 35 (13.4%) | 24 (25.5%) | 11(6.6%) | 0.108 (0.029 – 0.397) | 0.001 |
| **Sabaki** | 30 (11.5%) | 17 (18.1%) | 13 (7.8%) | 0.180 (0.049 – 0.665) | 0.010 |
| **Garashi** | 21 | 4 (4.3%) | 17 (10.2%) | **Ref** |  |
| Occupation |  |  |  |  | 0.473 |
| **Casual Labourer** | 10 (3.8%) | 3 (3.2%) | 7 (4.2%) | 1.287 (0.292 – 5.677) | 0.739 |
| **House wife** | 45 (17.2%) | 12 (12.8) % | 33 (19.8%) | 1.517 (0.617 – 3.730) | 0.364 |
| **Farmer** | 161 (61.7%) | 63 (67.0%) | 98 (58.7%) | 0.858 (0.432 – 1.707) | 0.663 |
| **Business** | 45 (17.2%) | 16 (17.0%) | 29 (17.4%) | **Ref** |  |
| Source of water |  |  |  |  | 0.505 |
| **Well and river** | 4 (1.5%) | 2 (2.1%) | 2 (1.2%) | 0.346 (0.042 – 2.831) | 0.322 |
| **Piped and pond** | 4 (1.5%) | 1 (1.1%) | 3 (1.8%) | 1.038 (0.095 – 11.296) | 0.975 |
| **Piped and river** | 23 (8.8%) | 11 (11.7%) | 12 (7.2%) | 0.378 (0.124 – 1.152) | 0.087 |
| **Pond** | 52 (19.9%) | 14 (14.9%) | 38 (22.8%) | 0.940 (0.354 – 2.490) | 0.900 |
| **River** | 121 (46.4%) | 48 (51.1%) | 73 (43.7%) | 0.526 (0.227 – 1.221) | 0.135 |
| **River and pond** | 2 (0.8%) | 1 (1.1%) | 1 (0.6%) | 0.346 (0.020 – 6.127) | 0.469 |
| **Well** | 20 (7.7%) | 8 (8.5%) | 12 (7.2%) | 0.519 (0.161 – 1.677) | 0.273 |
| **Piped** | 35 (13.4%) | 9 (9.6%) | 26 (15.6%) | **Ref** |  |
| Treatment for Bilharzia after diagnosis |  |  |  |  | 0.011 |
| **Received Treatment** | 38 (14.6%) | 20 (21.3%) | 18 (10.8%) | 0.422 (0.210 – 0.849) | 0.016 |
| **No treatment** | 7 (2.7) | 5 (5.3%) | 2 (1.2%) | 0.188 (0.036 – 0.992) | 0.049 |
| **No previous history** | 216 (82.8%) | 69 (73.4%) | 147 (88.0%) | **Ref** |  |
| Mode of contact with stagnant water |  |  |  |  | 0.110 |
| **When washing/fetching water** | 165 (63.2%) | 67 (71.3%) | 98 (58.7%) | 0.258 (0.073 – 0.916) | 0.036 |
| **When washing/fetching water and bathing** | 24 (9.2%) | 10 (10.6%) | 14 (8.4%) | 0.247 (0.057 – 1.076) | 0.063 |
| **When washing/fetching water and walking** | 4 (1.5%) | 1 (1.1%) | 3 (1.8%) | 0.529 (0.040 – 6.949) | 0.628 |
| **When bathing** | 31 (11.9%) | 11(11.7%) | 20 (12.0%) | 0.321 (0.777 – 1.342) | 0.119 |
| **When walking** | 17 (6.5%) | 2 (2.1%) | 15 (9.0%) | 1.324 (0.194 – 9.020) | 0.775 |
| **No contact** | 20 (7.7%) | 3 (3.2%) | 17 (10.2%) | **Ref** |  |
| Frequency of contact with stagnant water |  |  |  |  | 0.210 |
| **Less than twice a week** | 18 (6.9) | 8 (8.5%) | 10 (6.0%) | 0.221 (0.047 – 1.029) | 0.054 |
| **More than twice a week** | 172 (65.9) | 66 (70.2%) | 106 (63.5%) | 0.283 (0.080 – 1.004) | 0.051 |
| **Seasonally** | 51 (19.5) | 17 (18.1%) | 34 (20.4%) | 0.353 (0.091 – 1.373) | 0.133 |
| **No** | 20 (7.7) | 3 (3.2) | 17 (10.2) | **Ref** |  |
| Yes to bleeding outside menses;<br>approximately when |  |  |  |  | 0.017 |
| **Presently** | 11 (4.2%) | 10 (10.6%) | 1 (0.6%) | 0.047 (0.006 – 0.377) | <0.004 |
| **Within the last 6 months** | 12 (4.6%) | 5 (5.3%) | 7 (4.2%) | 0.663 (0.203 – 2.161) | 0.496 |
| **More than 6 months ago** | 14 (5.4) | 7 (4.2%) | 7 (4.2%) | 0.474 (0.160 – 1.401) | 0.177 |
| No | 224 (85.8%) | 72 (76.6%) | 152 (91.0%) | **Ref** |  |
| If yes to UTI, Cured or Not |  |  |  |  | 0.062 |
| **Yes** | 14 (5.4%) | 5 (5.3%) | 9 (5.4%) | 0.941 (0.305 – 2.900) | 0.915 |
| **Yes but recurred** | 11 (4.2%) | 8 (8.5%) | 3 (1.8%) | 0.196 (0.051 – 0.759) | 0.018 |
| **No UTI diagnosis** | 236 (90.4%) | 81 (86.2%) | 155 (92.8%) | **Ref** |  |
| Infection Intensity in urine<br>(eggs/10ml urine) |  |  |  |  | <0.001 |
| **<50 eggs/** | 25 (9.6%) | 20 (21.3%) | 5 (3.0%) | 0.104 (0.037 – 0.289) | <0.001 |
| **≥50** | 8 (3.1%) | 7 (7.4%) | 1(0.6%) | 0.059 (0.007 – 0.493) | 0.009 |
| **Negative** | 228 (87.4%) | 67 (71.3%) | 161 (96.4%) | **Ref** |  |

### Multivariable Logistic Regression Analysis for factors associated with FGS

Variables demonstrating a p-value < 0.25 in univariate analysis were included in the multivariable logistic regression model (**Table 12**). An alternative multivariable model that included urinary schistosomiasis positivity as a covariate is presented in Supplementary Table S1. In the adjusted model, infection intensity and sub-location of residence were independently associated with FGS. Urinary infection intensity was overall significant in the model (p < 0.001). Women with light infections (<50 eggs/10 ml) had elevated odds of FGS compared to women without detectable eggs (aOR 11.17, 95% CI 0.97–128.31; p = 0.053), although this estimate was borderline and imprecise. Heavy infections (≥50 eggs/10 ml) were not independently associated with FGS after adjustment (aOR 1.10; p = 0.947). Sub-location was significantly associated with FGS (p = 0.006). Participants residing in Mleji had more than sixfold higher odds of FGS compared to the reference area (aOR 6.30, 95% CI 1.96–20.22; p = 0.002), while those in Sabaki had over threefold increased odds (aOR 3.61, 95% CI 1.28–10.13; p = 0.015). Although Jimba demonstrated elevated odds (aOR 4.56; p = 0.083), this association did not reach statistical significance. Other sub-locations did not differ significantly from the reference. Age group, mode and frequency of water contact, history of urinary tract infection, and self-reported bleeding outside of menses were not independently associated with FGS in the final model (all p > 0.05).

**Table 12:**
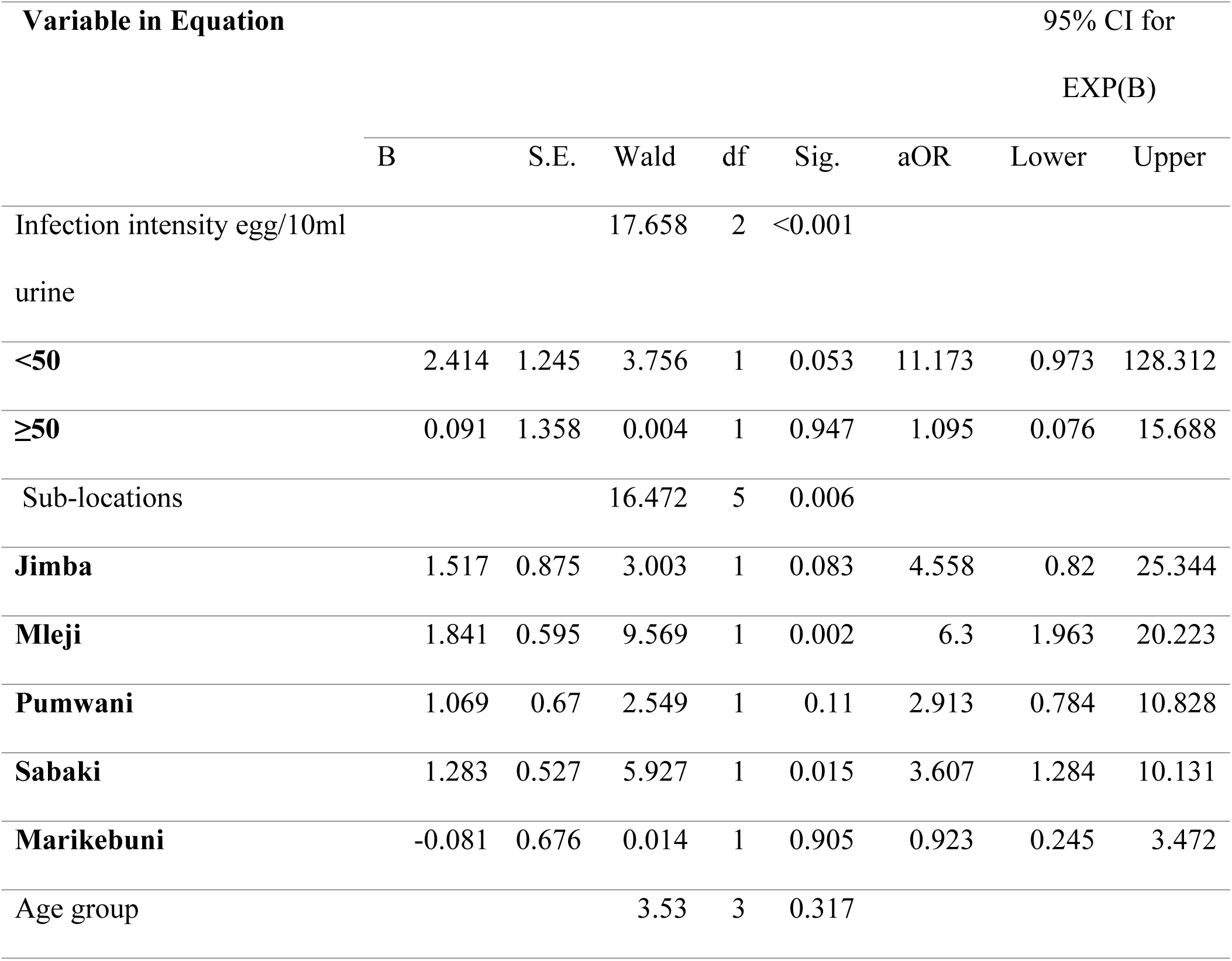
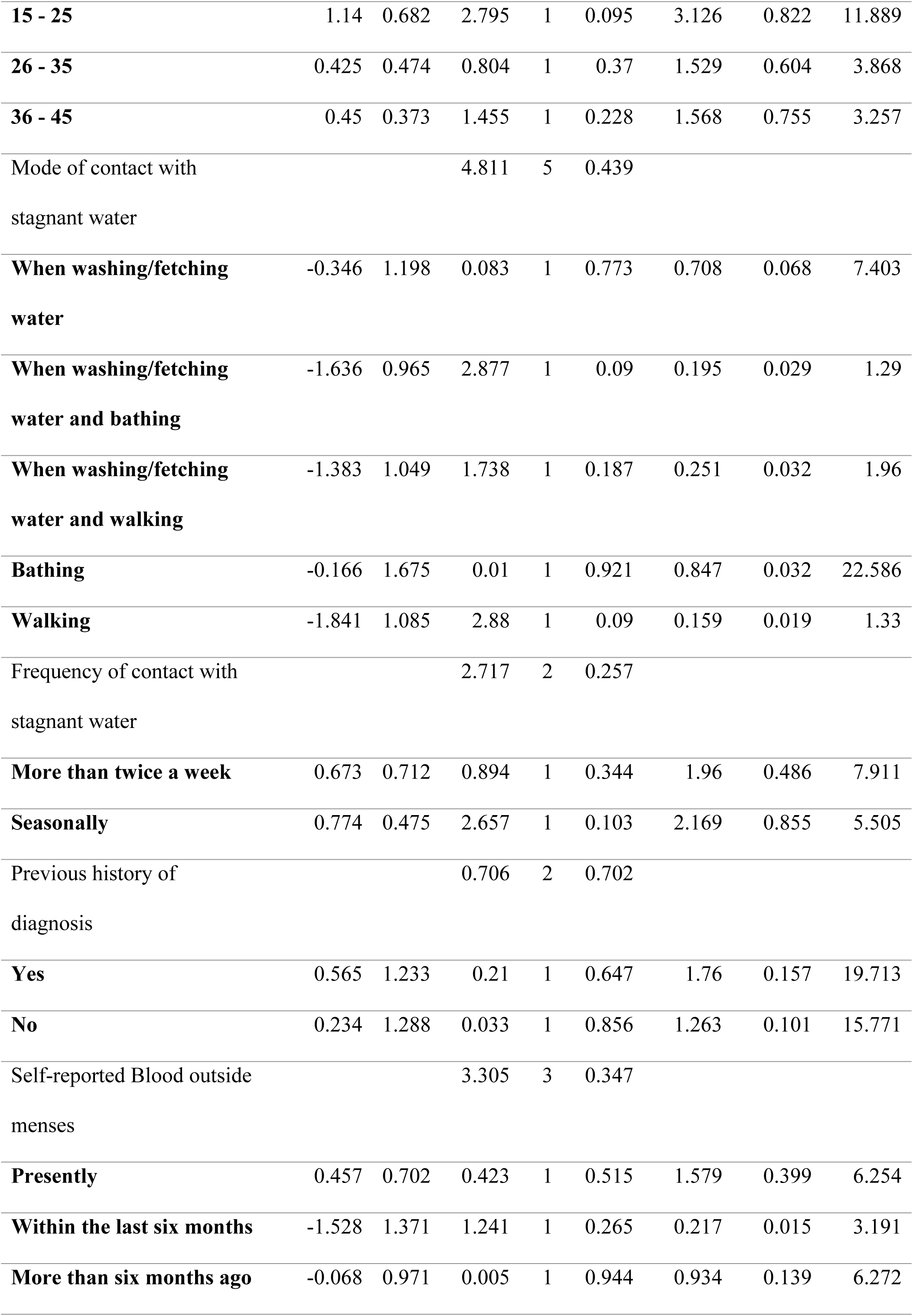
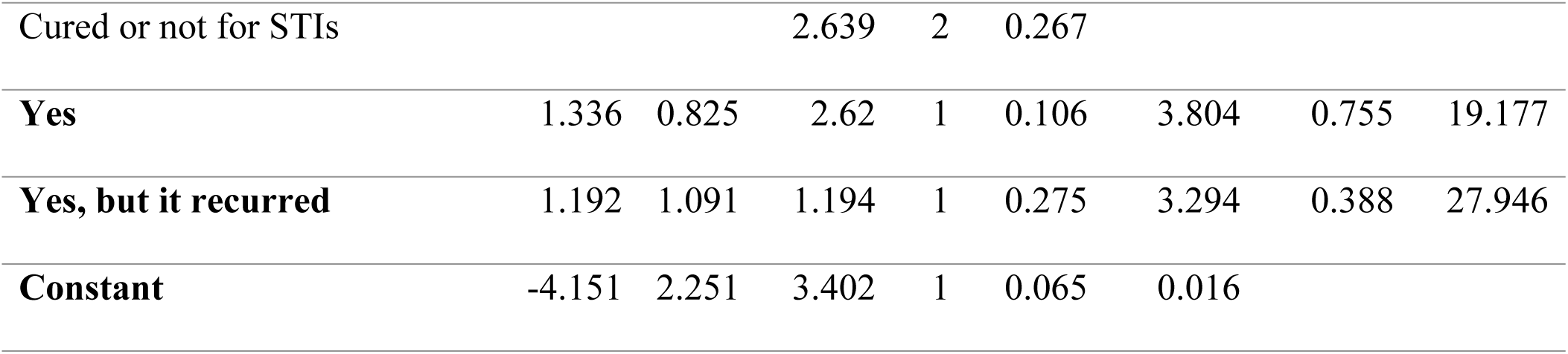
Adjusted Odds Ratios for Predictors of Female Genital Schistosomiasis in the Multivariate Logistic Regression Model, with a p-value less than 0.25.

## DISCUSSION

### Prevalence of Female Genital Schistosomiasis in Kilifi County

Despite the endemic transmission of *S. haematobium* along the Kenyan coast, population-based data on female genital schistosomiasis in adult women remain limited. This study demonstrates a high prevalence (36.0%) of PCR-confirmed female genital schistosomiasis (FGS) among women of reproductive age in Kilifi County. This estimate is consistent with reports from other *S. haematobium*–endemic settings in sub-Saharan Africa, where FGS prevalence ranges between 33% and 75% depending on the diagnostic approach and the population sampled (42,43). The prevalence observed here exceeds previously reported hospital-based data from neighbouring Kwale County (6), likely reflecting differences in study design and, importantly, the use of *Dra1*-based PCR for genital detection (34,35). Unlike urine microscopy, genital PCR directly detects schistosome DNA in reproductive tract specimens, improving sensitivity for detecting genital disease (34). The higher prevalence observed in this community-based study likely reflects improved detection rather than epidemiological divergence alone.

### Diagnostic Discordance between Urine Microscopy and Genital PCR

A key finding is that the majority of women with PCR-confirmed FGS had no detectable *S. haematobium* eggs in their urine. Although overall concordance between microscopy and PCR was 70.9%, positive agreement was limited to 26 concordant positive cases. This confirms significant under-detection of genital disease when relying on urine microscopy alone. Previous studies have similarly reported a poor correlation between urinary egg excretion and genital schistosomiasis (32,33). The biological explanation is plausible: adult worms reside in the pelvic venous plexus, and egg deposition may occur predominantly in genital tissues without consistent passage into the urinary tract (21,22). Furthermore, egg excretion is intermittent, reducing sensitivity even with repeated sampling (33). The non-linear association observed between PCR cycle threshold (Ct) values and mean egg counts further supports the concept that urinary parasite burden and genital DNA detection represent related but biologically distinct processes. The modest explanatory power of the polynomial model (R² = 9.6%) indicates that urinary egg intensity does not strongly predict genital DNA burden. Importantly, parasite DNA was detected in several women without urinary egg excretion, reinforcing the superior sensitivity of molecular diagnostics (34,44).

### Geographical Heterogeneity and Localised Risk

FGS prevalence differed significantly between sub-counties and sub-locations. Magarini sub-county had a higher burden compared to Rabai, and in multivariable analysis, sub-location remained independently associated with FGS (p = 0.006). Women residing in Mleji (aOR 6.30, p = 0.002) and Sabaki (aOR 3.61, p = 0.015) had significantly higher odds of infection compared with the reference area. Sabaki lies along the Athi–Galana–Sabaki River, which has previously been identified as a transmission hotspot with documented *Bulinus* spp snail habitats (45). Proximity to freshwater bodies is a well-established determinant of *S. haematobium* transmission (46–50). Although several behavioural exposures were associated with FGS in crude analyses, most did not remain independently significant after adjustment. This suggests that environmental and ecological clustering may play a more substantial role than individually reported behaviours in explaining geographic heterogeneity.

### Urinary Infection Intensity and Genital Involvement

Urinary infection intensity was overall significant in the multivariable model (p < 0.001). Women with low-intensity infections (<50 eggs/10 ml) had elevated odds of FGS compared to women without detectable eggs (aOR 11.17), although the association was borderline (p = 0.053) and imprecise, as indicated by the wide confidence interval. Heavy infection (≥50 eggs/10 ml) was not independently associated after adjustment. These findings suggest that genital involvement may occur even in low-intensity infections, consistent with evidence that tissue pathology is not strictly proportional to urinary egg burden (51). However, given the wide confidence intervals, these estimates should be interpreted cautiously and verified in larger studies.

### Clinical Manifestations and Symptom Correlates

In bivariate analyses, haematuria, dysuria, bleeding outside menses, and urinary egg positivity were significantly associated with FGS. Haematuria has long been recognised as a proxy marker for urogenital schistosomiasis (52–54). However, haematuria did not remain independently associated in the adjusted model, indicating that while it reflects urinary infection, it cannot reliably predict genital involvement. Bleeding outside menses was more common among women with FGS in unadjusted analyses. Chronic inflammation, mucosal disruption, and angiogenesis induced by egg deposition may explain abnormal genital bleeding (1,11,51). However, this variable was self-reported and not clinically verified. Moreover, it did not retain significance in multivariable modelling. Similarly, dysuria and a history of recurrent UTIs did not remain independently associated after adjustment. These findings suggest that symptom-based screening alone is likely to lack specificity for identifying FGS in endemic communities.

### Age Distribution and Exposure Patterns

FGS prevalence varied across different age groups, with the highest crude prevalence observed among women aged 15–25 years. Similar patterns have been reported in other endemic settings, which may indicate early and repeated exposure to water starting in adolescence (55). However, the age group did not remain independently associated in the adjusted model. Although egg counts increased descriptively with age, no statistically significant correlation was found. These findings suggest that age is not an independent predictor of genital disease in this cohort.

### Implications for Surveillance and Control

Historically, schistosomiasis control strategies in Kenya have focused on school-aged children through school-based preventive chemotherapy. The World Health Organisation (WHO) now recommends that preventive chemotherapy also be administered to at-risk adults in endemic settings, particularly in moderate- and high-transmission areas (56). However, the implementation of coverage, frequency, and adult participation can vary across different transmission zones, and preventive chemotherapy does not specifically address the detection of genital pathology. In this study, nearly three-quarters of women with PCR-confirmed FGS had no detectable urinary eggs. This suggests that genital disease may go unrecognised when surveillance relies mainly on urine microscopy or presumptive mass drug administration without a genital assessment. While expanding preventive chemotherapy is crucial for reducing transmission intensity, it may not effectively identify women with established genital involvement. Integrating genital self-sampling and molecular diagnostics into reproductive health services could enhance case detection in high-burden areas. However, the feasibility, cost-effectiveness, and scalability of these approaches require further evaluation within routine health systems. Strengthening water, sanitation, and hygiene (WASH) interventions while targeting high-risk localities such as Sabaki and Mleji may further reduce transmission intensity and long-term reproductive morbidity.

### Study Limitations

This study has several limitations. The cross-sectional design prevents causal inference. Attrition throughout the study phases reduced the analytic sample from 336 women enrolled for urinary screening to 261 who provided complete data for urine microscopy, genital PCR, and questionnaires. Additionally, some genital specimens did not meet quality control standards. These factors may have introduced selection bias, making prevalence estimates potentially not fully generalisable to all women initially enrolled. Some multivariable estimates were imprecise, as indicated by wide confidence intervals, likely due to small sample sizes in certain exposure categories. Clinical symptoms were self-reported, which may have introduced recall and reporting bias. Colposcopic examination, a standard method for identifying characteristic FGS lesions, was not performed. Consequently, the clinicopathological correlation between molecular detection and the morphology of genital lesions could not be assessed.

## CONCLUSION

Community-based data on female genital schistosomiasis (FGS) among women of reproductive age in coastal Kenya are still limited. This study employed molecular testing and found a high prevalence of FGS (36.0%) in Kilifi County, with 72% of affected women showing no detectable *S. haematobium* eggs in their urine. These results suggest that relying solely on urine microscopy may significantly underestimate genital involvement. To improve the identification of FGS in endemic areas, it is essential to incorporate genital sampling and molecular assays into surveillance and case detection strategies. Additionally, strengthening water, sanitation, and hygiene (WASH) interventions, along with ensuring adequate preventive chemotherapy coverage for at-risk populations, may further reduce transmission intensity and long-term reproductive morbidity.

## Data Availability

All relevant data supporting the findings of this study are provided within the manuscript and its Supporting Information files.

## Acknowledgments

We sincerely thank all the women who participated in this study for their time and willingness to contribute. We are grateful to the Kilifi County Department of Health and the sub-county health management teams for their support and collaboration during field activities. We acknowledge the assistance of local health facility staff and community health volunteers who facilitated participant mobilisation and sample collection. We also acknowledge the Centre for Molecular Biosciences and Genomics (CMB Genomics) Laboratory for their technical support with molecular analysis, particularly in processing and detecting *Schistosoma haematobium* DNA from genital samples. The first author recognises support from the Organisation for Women in Science for the Developing World (OWSD) through the Sandwich Programme, which supported her research training. This support did not influence the study design, data collection, analysis, or interpretation. Finally, we appreciate the guidance and mentorship provided by our supervisors and collaborators throughout the study.

